# Anticholinergic drugs and clinical outcomes in older people with and without dementia- A systematic Review

**DOI:** 10.1101/2025.03.14.25323976

**Authors:** Delia Bishara, Katrina Davis, Christoph Mueller, Olubanke Dzahini, Nicola Funnell, Justin Sauer, Daniel Harwood, David Taylor, Robert Stewart

## Abstract

**Background:** Anticholinergic medications are widely used, however their use in older people has been linked to cognitive decline, dementia and increased mortality. This systematic review examines the literature investigating relationships between anticholinergic burden and risk of dementia, cognitive impairment, and outcomes in dementia.

**Methods:** Cochrane database and PubMed searches using the terms “anticholinergic” and “dementia” or “cognition” were performed up to May 2023. Outcomes included: (i) dementia diagnosis, (ii) cognitive outcomes in people without dementia (iii) cognitive outcomes, hospitalisation and death in people with dementia. Inclusion and exclusion criteria were defined, and papers were evaluated for inclusion by two researchers independently. Papers examining these relationships specifically for urinary drugs and antidepressants were also analysed separately.

**Results:** Sixty observational studies met our criteria across the three outcomes of interest. Anticholinergic burden was found to be consistently associated with increased risk of dementia however the relationships with cognitive outcomes were less clear. In people with dementia, there were consistent associations between the anticholinergic burden and mortality (hazard ratio (HR) range: 1.04-1.23) or hospitalisation (HR range: 1.13-4.54) but not for cognitive outcomes. Urological drugs with high anticholinergic burden were associated with a ≥50% increased mortality risk in people with dementia.

**Conclusion:** Anticholinergic burden has been consistently associated with increased dementia incidence. Furthermore, in people with existing dementia, anticholinergic burden is associated with increased mortality and hospitalisation. Associations with cognitive outcomes in people without/with dementia remains uncertain. Clinicians should be advised to exercise caution with anticholinergic medication use in older people.

## Introduction

Anticholinergic medications are widely used to treat many medical conditions, however they have been linked to cognitive decline^1^, dementia and higher mortality^2 3^. Drugs for urinary frequency and antidepressants have also been linked with a higher risk of dementia^2 3^. Our objective was to review associations between anticholinergic agents in older people and associations with cognition, dementia risk, and dementia clinical outcomes.

## Methods

Cochrane database and PubMed searches were executed on 4^th^February 2020 using the keywords “anticholinergic” and “dementia” or “cognition”. The search was limited to English language, “human” subjects and to publications within the last 10 years. Three top-up searches were made to bring date of inclusion to articles indexed before May 2023. Abstracts were examined to identify original research and reviews comparing groups of older people (>50 years) with or without dementia with different anticholinergic drug load. We identified three related groups for participants and outcomes: Group 1. Risk of dementia, Group 2. Cognitive outcomes in people without dementia, and Group 3. Outcomes in people with dementia. Our inclusion criteria are described below.

*Inclusion criteria for papers examining the associations between anticholinergic agents and:*

– (Group 1) dementia
– (Group 2) cognitive outcomes
– (Group 3) outcomes in dementia: mortality, hospitalisation or cognitive outcomes:

– **Population**- Recruitment from any setting. Group 1 and 2: Older adults >50 years without cognitive impairment. Group 3: Older adults >50 years with dementia.
– **Index prognostic factor-** Groups with difference in anticholinergic burden (using validated scale).
– **Comparative prognostic factors (covariates of interest)-** Studies must report, and preferably control for, baseline age, gender, comorbidities (e.g., using Charlson Comorbidity Index) and severity of dementia (where relevant) by Mini Mental State Examination (MMSE) or other cognitive measure.
– **Outcomes-**

Group 1: Diagnosis of dementia.
Group 2: Outcome from standardised cognitive assessments (e.g., MMSE).
Group 3: Outcome from standardised cognitive assessments, hospitalisation, mortality.
– **Study:** Any design. Minimum size, 10 participants.

Studies comprising of 10 subjects or fewer were excluded as were studies including some subjects younger than 50 years old. Where systematic reviews were available, these took precedence over original research.

The PRISMA flow chart (see Figure 1) describes the selection process. DB and KD examined the abstracts and full papers independently to determine whether they met inclusion criteria with discussion and resolution of any disagreement. The search was updated three times to include new papers using identical selection criteria.

**Figure 1.**
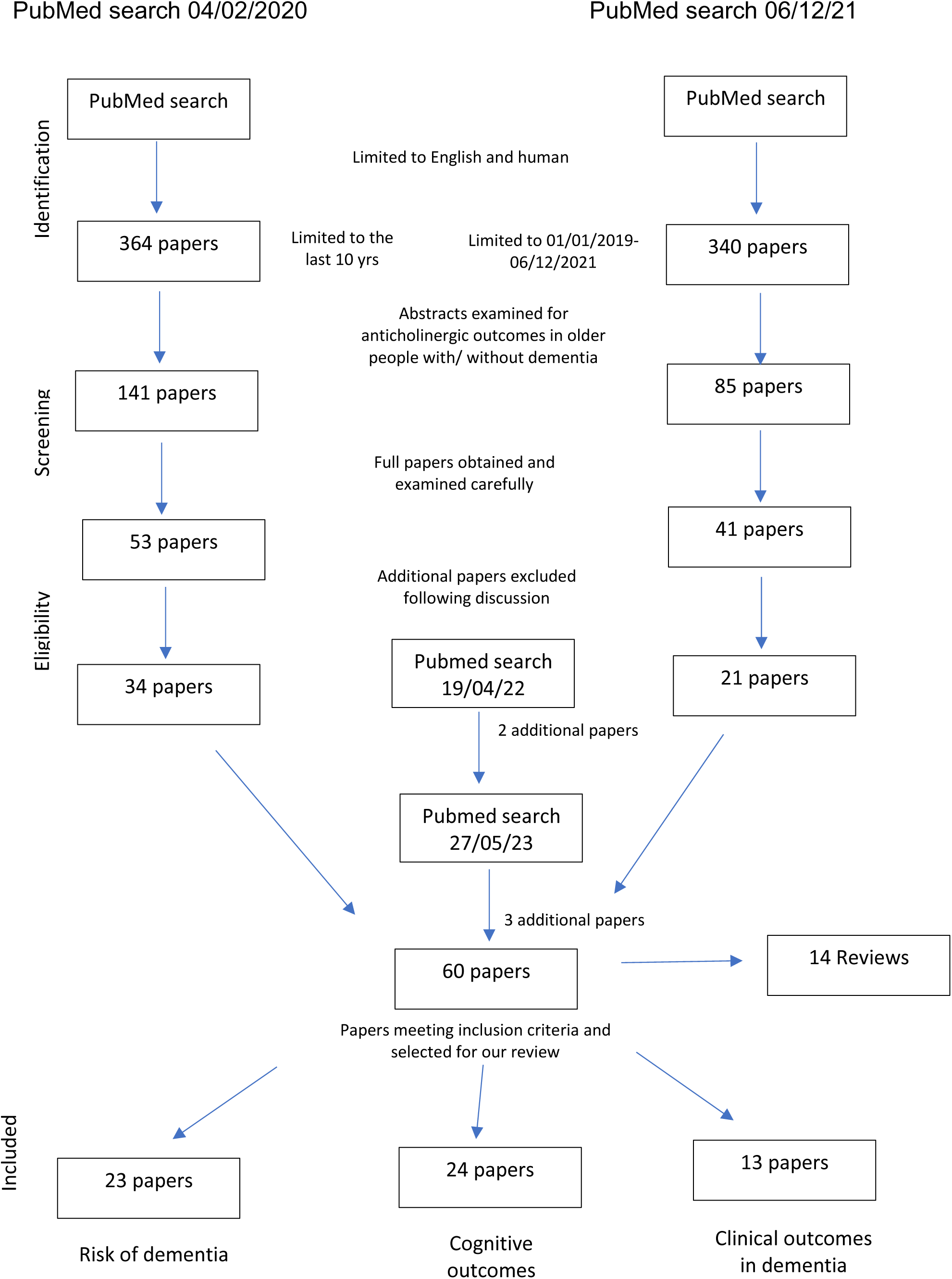
Prisma flow diagram of the study selection process

Where required, we contacted authors for additional information. Exclusions were primarily due to the wrong or unreported population, no outcome of interest or lack of covariate reporting. Several otherwise eligible studies were excluded because a recognised anticholinergic burden scale had not been used. An exception was made for within-class comparisons of antidepressants and drugs used in urinary conditions, which we retained and analysed separately. The rational for this was that an anticholinergic burden scale was not deemed necessary where individual drugs were directly compared against others within the same class.

We carried out back-referencing of reviews and papers, and forward-referencing using Google Scholar, to ascertain any missed studies meeting inclusion criteria. Study quality was assessed using the Newcastle-Ottawa Scale, separately rated by DM and KD resulting in an agreed consensus score. “Selection” has four items on the definition of cohort and exposure. “Comparability” has two items regarding control of major confounders (age, sex and comorbidities), and “Outcome” has three items regarding the definition and ascertainment of outcome, including whether exposure and observation were long enough to see the plausible effect. There is no agreed definition as to how to combine these scores; we present the sum for reference.

### Analysis

Endnote was used to organise references. MS Excel was used to collect and organise extracted information. The Cochrane Foundation Review Manager (RevMan version 5.4) was used to draw plots. We planned to carry out meta-analysis if there was a group of more than two studies that had compatible exposure and outcome measures for combining.

## Results

Sixty papers met inclusion criteria for at least one outcome of interest. Twenty-four investigated cognitive outcomes in people without dementia, 23 the risk of incident dementia, and 13 outcomes in dementia. Thirteen reviews in groups 1 and 2 were identified and summarised. Only one review of outcomes in dementia (group 3) was identified. Due to the limited evidence in this area, this review focussed mainly on dementia outcomes; however study data for dementia risk and cognitive outcomes can be found in the supplementary material.

### Anticholinergic burden and risk of dementia

Twenty-three studies were included, comprising over 3 million patients across the US, Europe, Australia and Asia (Supplementary Table 1). All were observational, including longitudinal database studies, conventional cohort and case-control studies. A significant association between anticholinergic drug use and dementia risk was observed in most studies. Systematic reviews confirmed these findings. One systematic review and meta-analysis concluded that anticholinergic use for ≥ 3 months increased the risk of dementia on average by an estimated 46%^5^, a dose-dependent relationship concluded by another^6^. Both reviews highlighted antidepressants, antiparkinsonian and urological drugs as associated with higher risk.

### Anticholinergic burden and cognitive outcomes

Twenty-four studies of nearly 150,000 patients investigated the effects of anticholinergic agents on cognitive outcomes (Supplementary Table 2). Studies were mostly conducted in the USA, Europe, and Australia with one in Canada and one in Taiwan; they included observational research, both cross-sectional and longitudinal. Fifteen reported a significant association between anticholinergic burden and cognitive outcomes, despite different study design, anticholinergic burden scales, populations, and cognitive measures. Of the remainder, four reported mixed results and five studies found no significant association. Three systematic reviews^7–9^ concluded an association between anticholinergic medication and significant decline in cognition, although this was not observed for all studies.

### Anticholinergic burden and mortality, hospitalisation or cognitive outcomes in people with dementia

Thirteen studies from our review met inclusion criteria (Table 1). Further details of quality assessments are shown in Table 2. No paper scored full points on the quality scale, but many dropped only one mark, for selecting only people with dementia who were on medication or attending a specialist clinic, thereby reducing generalisability.

**Table 1.** Papers examining the association between anticholinergic burden and cognitive decline, hospitalisation or mortality in people with dementia.

| Authors/<br>reference | Study Design | Subjects | Sample<br>size/<br>Nation | Anti-<br>cholinergic<br>scale used | Aim/ Main outcomes | Significant<br>association | Outcome for<br>meta-analysis |
| --- | --- | --- | --- | --- | --- | --- | --- |
| Ah 2019 <sup>10</sup> | Retrospective cohort study. Events for mortality followed up for 2 years after index date | Newly diagnosed older adult (>60 yrs) dementia patients. Median age (IQR): 60-74 yrs 2902 (39.0) 75-84 yrs 3789 (50.9) ≥85 yrs 747 (10.0) | N= 25, 825<br>Korea | ACB | Evaluation of anticholinergic burden on dementia treatment modification, delirium and mortality. In patients with high anticholinergic burden (ACB >3), significantly more patients experienced treatment modification (34.9% vs 32.1%), delirium (5.6% vs 3.6%) or mortality (16.8% vs 14.1%) than controls. ACB score >3 within the first 3 months significantly increased the risk of treatment modification (HR: 1.12, CI: 1.02-1.24), delirium (HR: 1.52, CI: 1.17-1.96) and mortality (HR: 1.23, 95% CI:1.06-1.41). | Yes | Mortality:<br>(HR: 1.23, 95% CI:1.06-1.41)<br>No data for hospitalisation |
| Bishara 2020 <sup>11</sup> | Retrospective cohort study | Patients ≥ 50 yrs, diagnosed with dementia. Mean age 79.8 yrs. | N= 14, 093<br>UK | AEC | To investigate associations between central AEC and mortality, hospitalisation and cognitive decline in patients with dementia. Patients with AEC score ≥ 2 or total AEC score ≥ 3 had an increased risk of mortality (hazard ratio 1.07; 95% CI: 1.01-1.15) and hospitalisation (1.10; 95% CI: 1.04-1.17), but had primarily, acutely impaired cognitive function, rather than longer-term differences in cognitive decline. Finally, a 1-point increase in AEC score was associated with a 2% increased risk of mortality indicating a dose-response relationship. | Yes for mortality and hospitalisation<br>No for cognitive decline | Mortality:<br>(HR: 1.10, 95% CI:1.03-1.18)<br>Hospitalisation:<br>(HR: 1.13, 95% CI:1.07-1.21) |
| Buckley 2022<sup>15</sup> | Retrospective registry-based observational cohort study | Patients with MCI or dementia presenting to a tertiary-referral memory clinic from 2013 to 2019 Median age 79 years. | N= 418<br>N=261 dementia, N=157 MCI)<br>Dublin, Ireland | ACB | Investigating the association between potentially inappropriate medicines related to cognitive impairment and anticholinergic burden with mortality in dementia or mild cognitive impairment (MCI).<br>ACB≥3 All-cause death 0.87 (0.46–1.64) p=0.67 | No for all-cause mortality. | Mortality:<br>(HR: 0.87, 95% CI: 0.46-1.64) p=0.67 |
| Cross 2017 <sup>14</sup> | Cross-sectional longitudinal (3 yrs) analyses of data from Prospective Research In MEemory clinics (PRIME) study. | Community-dwelling people who attended memory clinics and had a diagnosis of MCI or dementia. Mean age (SD) 77.6 yrs (7.4). | Total N= 964<br>N=384 people with dementia completed study | ACB | To examine if potentially inappropriate medicines in people with cognitive impairment and anticholinergic cognitive burden were associated with mortality. ACB scores (adjusted hazard ratio: 1.18 95% CI: 1.06–1.32) were associated with mortality. | Yes | Mortality:<br>HR per 1-unit ACB score<br>(HR: 1.15, 95% CI:1.01–1.31)<br>No data for hospitalisation |
|  |  |  | Australia |  | For people with dementia only ACB was associated with mortality in adjusted model (adjusted hazard ratio: 2.08 95% CI: 0.98–4.42) |  |  |
| Dyer 2020 <sup>21</sup> | Analysis of data from NILVAD, trial of Nilvadipine in AD. Longitudinal observational study. | Patients ≥ 50 yrs, diagnosed with AD. Mean age was 77.88 (±8.25) yrs. | N=142<br>Ireland | ACB | To assess if anticholinergics are associated with accelerated cognitive decline in mild/ moderate AD. ACB score was not associated with greater progression on the ADAS-Cog/CDR-sb over time, but a higher total ACB predicted greater dementia severity on the Disability Assessment for Dementia (DAD), which persisted after robust covariate adjustment (β Coef: -1.53, 95% CI: -2.83 to -0.23, p= .021). There was a significant interaction between APOE ε4 status and ACB score, with carriers experiencing greater progression on CDR-Sb (β Coef: 0.36, 95% CI: 0.05–0.67, p= .021) and DAD (β Coef: -3.84, 95% CI: -7.65 to 0.03, p= .049) | No for cognitive decline<br>Yes for dementia severity | No suitable results for meta-analysis |
| Fox 2011 <sup>22</sup> | Study is part of a larger longitudinal cohort study (LASER-AD) | Participants with AD, mean age 81.0 years [SD 7.4 (range 55-98)] | N=224<br>UK | ABS | To examine the effect of anticholinergics on cognitive impairment and deterioration in AD. There were no differences in MMSE and other cognitive functioning at either 6 or 18 months after adjusting for covariates between those with and those without high anticholinergic load. | No for cognitive impairment and deterioration in AD | No suitable results for meta-analysis |
| Gnjidic 2014 <sup>12</sup> | Retrospective cohort study. Data extracted from various registers. | Community-dwelling people aged 65 years and over with and without AD. Mean age of people with and without AD was 79.2 years. There were more women (66.5%) than men. | N=16,603 with dementia<br>N=16,603 matched<br>Finland | DBI | Relationship between cumulative exposure of anticholinergics and sedatives and hospitalisation/ mortality in people with/ without AD. Among people with AD, for every unit increase in DBI (equivalent to exposure to 2 or more anticholinergics and sedatives at minimum effective dose), adjusted HR for mortality was 1.21 (95%CI: 1.09–1.33). When using no DBI exposure as reference group, adjusted HR for mortality for high DBI group (≥1) was 1.34 (95%CI: 1.13– 1.60). For every unit increase in DBI, adjusted IRR for length of hospital stay in people with AD was 1.06 (95%CI: 0.99– 1.12). When using no DBI exposure as the reference group, adjusted IRR for high DBI group (≥1) was 1.15 (95%CI: 1.05–1.26). The number of separate hospital admissions increased with higher DBI exposure, with an adjusted IRR of 1.22 (95%CI: 1.17– 1.27). There was a dose-response relationship between higher DBI exposure and higher number of separate hospital admissions in people with and without AD | Yes for mortality and hospitalisation | Mortality:<br>(HR: 1.21 (95%CI, 1.09-1.33)<br>Hospitalisation:<br>(Adjusted IRR:1.22 (95% CI:1.17–1.27) |
| Lampela 2013 <sup>23</sup> | Data analysed from population-based Geriatric Multidisciplinary Good Care of the Elderly Study | Randomly selected persons aged 75 years and over assigned to intervention. Mean age (SD) 81.7 (4.9) | N= 621<br>N=129 with dementia and N=492 without dementia.<br>Finland | SAA assay, Carnahan's Anticholinergic Drug Scale, Rudolph's Anticholinergic Risk Scale & Chew's list. | Study investigated whether SAA assay results and scores from 3 ranked lists of anticholinergic drugs are associated with anticholinergic adverse events in older people.<br>No association was found with anticholinergic scores and MMSE in people with dementia. | No for cognitive deterioration | No suitable results for meta-analysis |
| Liang 2023<sup>20</sup> | Retrospective cohort study. Analysis of data collected from hospital electronic health records. | Patients aged ≥65 years admitted to an internal medicine ward. | N=3061<br>N=284 with dementia<br>N=2,777 without dementia | ACB | Examining the association between ACB scores with readmission and emergency room (ER) revisit after discharge.<br>In those with dementia, only major polypharmacy, but not ACB scores, correlated significantly with readmission and ER revisit, except readmission within 3 months. No significant association between ACB scores and readmission or ER visit for dementia patients within 6 months (odds ratio OR: 1.02, CI: 0.80-1.30) and (OR: 1.11, CI: 0.88-1.42) | No for admission | Admission within 6 months: (OR: 1.02; CI: 0.80-1.30) |
| McMichael 2021 <sup>4</sup> | Longitudinal observational study (up to 6yrs). | Patients prescribed ≥1 dementia medication from prescribing database. Mean age 77.2 (SD± 8.3) yrs. | N=25,418<br>Northern Ireland | ACB | Influence of anticholinergic burden on mortality rates in people with dementia.<br>Higher anticholinergic burden was associated with significantly higher mortality rates in comparison to people with dementia who had no anticholinergic burden in unadjusted models (HR: 1.59, 95% CI:1.07-2.36). Urological (HR:1.20, 95% CI:1.05-1.38) and respiratory (HR: 1.17, 95% CI:1.08-1.27) drugs significantly increased mortality rates. | Yes for mortality | Mortality: Unadjusted model: (HR: 1.59: 95% CI: 1.07-2.36) Adjusted model: (HR: 1.17: 95% CI: 1.11–1.24)<br>No data for hospitalisation |
| Sanders 2017 <sup>24</sup> | Data from 2 studies: a cross-sectional study and baseline data from related Dutch intervention study | Nursing home residents > 70 yrs with dementia. Mean age (SD) 85.13 (5.69) yrs. | N= 140<br>N= 74 and N= 66<br>The Netherlands | DBI | Determining the relationship between DBI and cognitive and physical functions.<br>Multivariate models yielded no associations between total drug burden (TDB), anticholinergic DBI, and sedative DBI, and physical or cognitive function (MMSE) (all p > 0.05). | No for physical or cognitive function | No suitable results for meta-analysis |
| Tan 2018 <sup>13</sup> | Cohort study of people registered in the Swedish Dementia Registry from 2008-2014. Mean follow-up of 2.31 (SD 1.66) yrs | People with dementia and no prior history of stroke. Mean age of 79.9 (SD, 7.90) yrs with the majority being female (60.7%). | N= 39, 107<br>Sweden | ACB | Association between total anticholinergic cognitive burden and risk of stroke and death.<br>During follow-up 11,224 (28.7%) individuals had a stroke or died. Compared with non-users of anticholinergics, ACB score of 1 (HR 1.09, 95% CI 1.04-1.14) and ACB score of ≥2 (HR 1.20, 95% CI 1.14-1.26) increased the risk of developing stroke and death, indicating a dose-response relationship. | Yes for stroke and death | Death (continuous): (HR: 1.04, 95% CI:1.02-1.06) |
|  |  |  |  |  |  |  | No data for hospitalisation |
| Watanabe 2018 <sup>19</sup> | Retrospective chart-based study (mean follow-up period= 420±366 days) | Outpatients with a diagnosis of dementia, mean age 78±7 years | N= 61<br>Japan | ARS | Investigation of the influence of anticholinergic medication on the risk of hospitalisation. Anticholinergic medication was related to a higher risk for hospitalisation (crude hazard ratio: 3.62, 95% confidence interval: 1.45–9.04, *P*<0.01; adjusted hazard ratio: 4.54, 95% confidence interval: 1.03–20.0, *P*<0.05). In contrast, Mini-Mental State Examination score, Charlson Comorbidity Index, and the number of drugs were not major risk factors for hospitalisation in patients with dementia. | Yes for hospitalisation | No data for mortality<br>Hospitalisation: (HR: 4.54, 95% CI: 1.03–20.0) |

ACB= Anticholinergic cognitive burden scale ABS= Anticholinergic Burden Score ADS= Anticholinergic Drug Scale AEC= Anticholinergic Effect on Cognition Scale ARS= Anticholinergic Risk Scale DBI= Drug Burden Index AD= Alzheimer's Disease MCI= Mild Cognitive Impairment

**Table 2.**
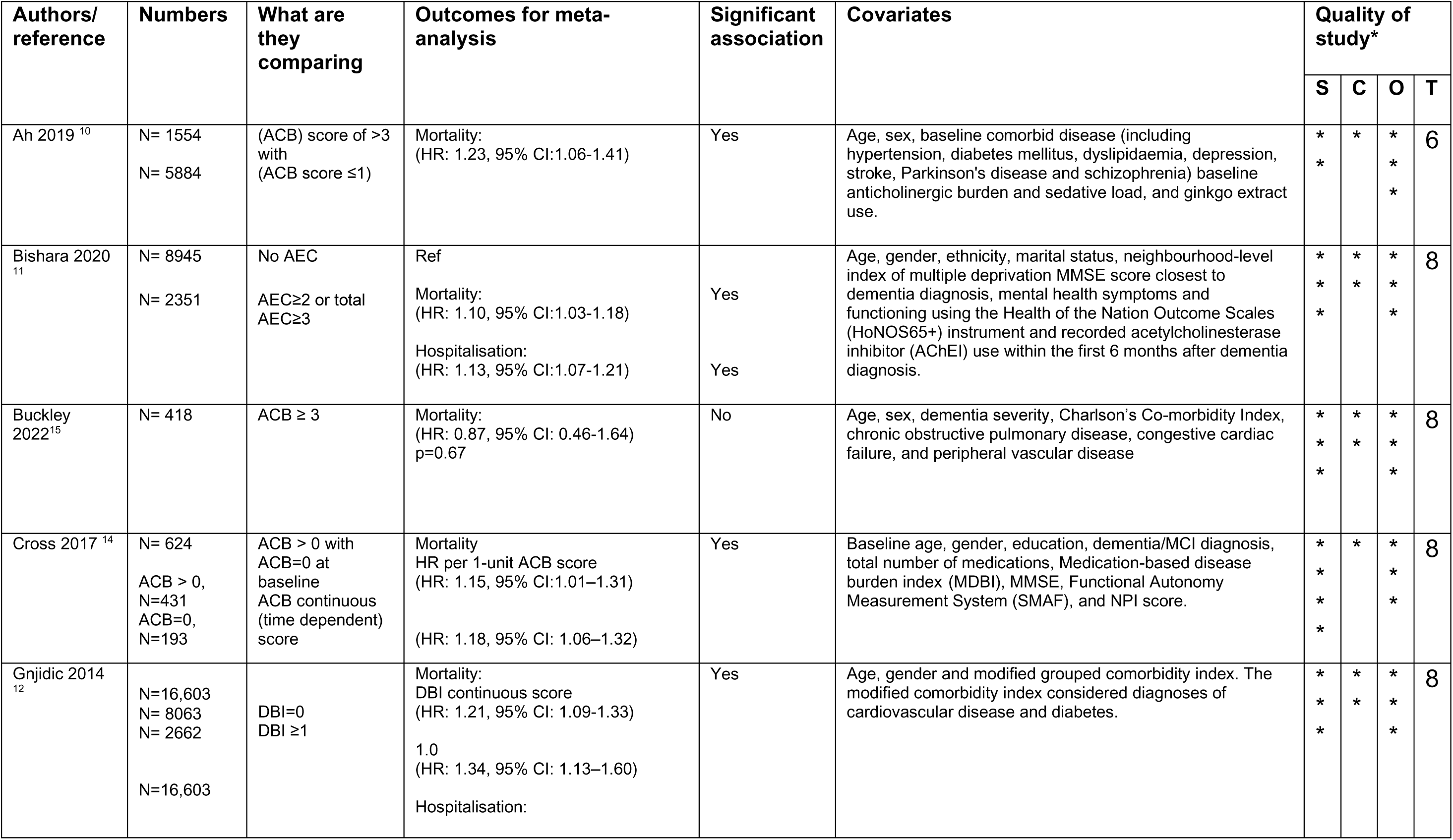
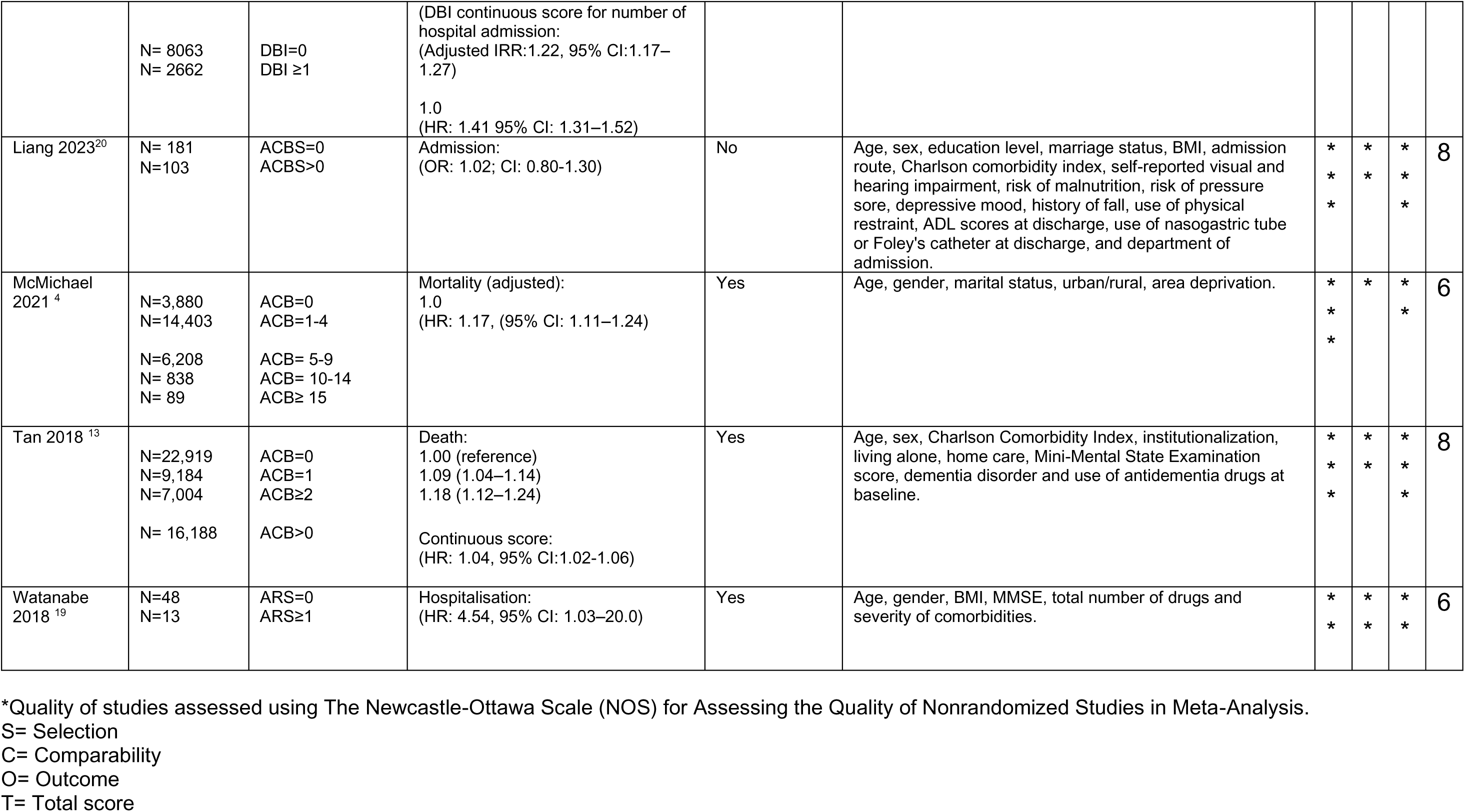
Summary of study outcomes for papers (included in meta-analysis) examining the association between anticholinergic burden and cognitive decline, hospitalisation or mortality in people with dementia.

#### Anticholinergic burden and mortality in dementia

Table 1 describes seven studies^4 10–15^ with mortality as an outcome, all of which were longitudinal cohort studies, five of which included over 10,000 participants. Six of the seven studies found a significant effect on mortality, with only one^15^ showing no association; however, this included patients with both Mild Cognitive Impairment and dementia.

Combining data from different studies was hampered by differences in anticholinergic burden scale used and the anticholinergic burden scores that were compared, except for two studies^14 16^. The forest plot (Figure 2a) illustrates a consistent significant excess risk of death associated with a higher anticholinergic burden of effect size ranging between HR of 1.04 to 1.23. Moreover, three^11–13^ of seven studies showed a dose-response relationship. Three^11 12 17^ rated highly on the NOS (concerns were about representativeness only), while two had other quality concerns^10 4^. Considering noteworthy secondary findings, elevated mortality was associated with urological and respiratory anticholinergic drugs, but not with antidepressants^4^. Also the association between ACB and mortality was consistent across dementia sub-types^13^. Only one relevant review was identified^18^ which also concluded an association between anticholinergic medication and all-cause mortality.

**Figure 2a).**
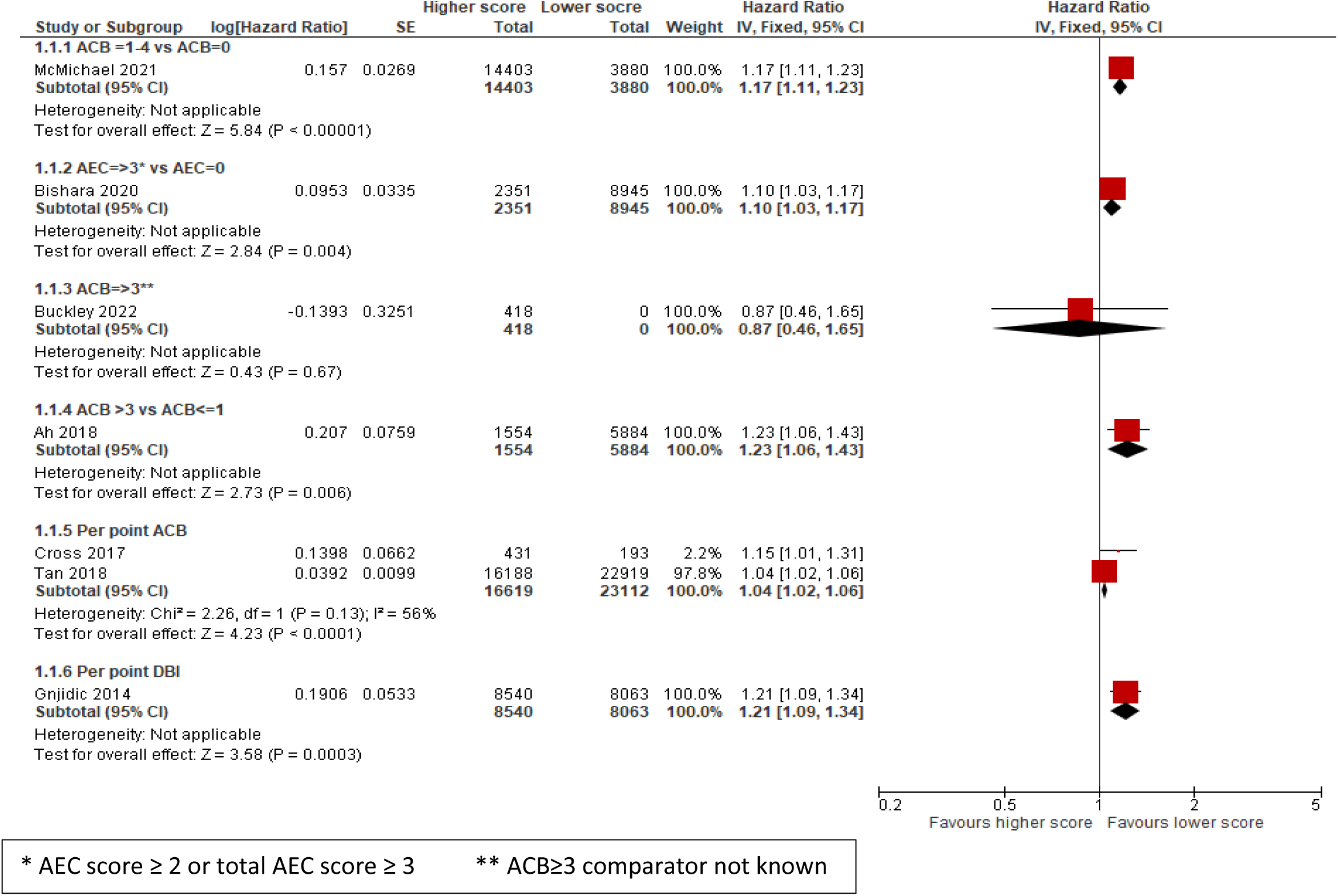
**Forest plot of hazard ratios for mortality**

#### Anticholinergic burden and hospitalisation in dementia

Of four studies^11 12 19 20^ examining the association between anticholinergic burden and hospitalisation in people with dementia (Table 1), three^11 12 19^ found a higher risk and one^20^ found no association. In NOS assessment, quality concerns for three^11 12 20^ were for representativeness only, while one^19^ had some other quality concerns. Again, a meta-analysis could not be carried out as each study used a different exposure scale. The forest plot (Figure 2b) illustrates a higher anticholinergic burden associated with significantly more hospitalisation with HRs ranging from 1.13 to 4.54. The review by Wang^18^ also noted longer hospital length of stay in three studies.

**Figure 2b).**
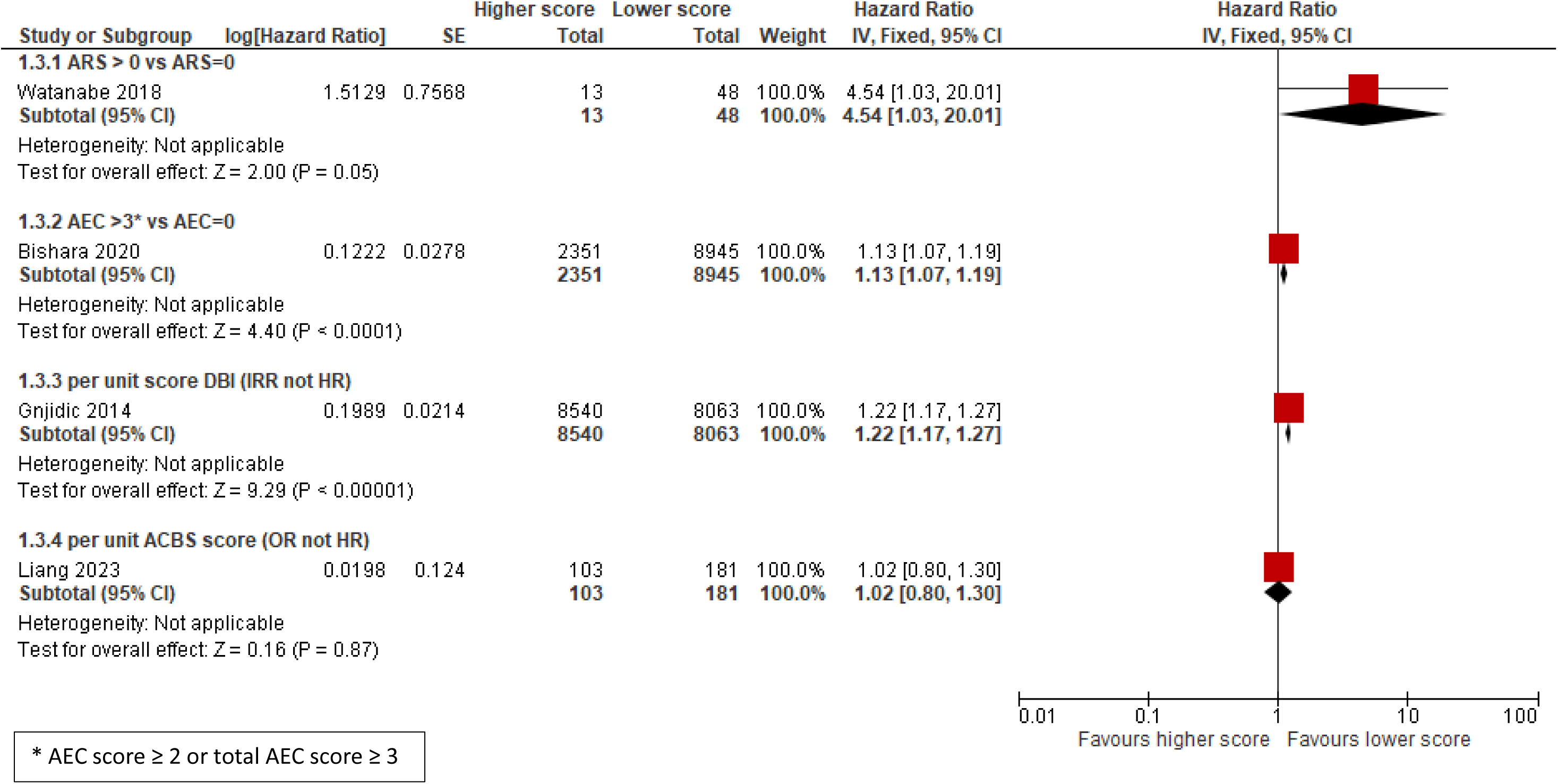
**Forest plot of hazard ratios for hospitalisation**

#### Anticholinergic burden and cognitive outcomes in dementia

Of the 13 studies of dementia cohorts, 5 examined cognitive outcomes in dementia and none found an association with anticholinergic burden (Table 1). Bishara 2020^11^ found that anticholinergic burden was associated with impaired cognitive function around the dementia diagnosis, rather than differences in subsequent cognitive decline; Dyer 2020^21^ found that anticholinergic burden was not associated with greater progression on the ADAS-Cog/CDR-sb over time, although a higher burden predicted greater dementia severity which persisted after robust covariate adjustment. Fox 2011^22^ found no differences in MMSE or other cognitive functioning at either 6 or 18 months, after adjusting for covariates, between those with and without high anticholinergic load. Similarly, Lampela 2013^23^ found no association between anticholinergic scores and MMSE in people with dementia, and Sanders 2017^24^ also found no associations in multivariate models for total drug burden, anticholinergic burden, or sedative burden with physical or cognitive function. The review by Wang^18^ concluded inconsistent results due to varying study characteristics and quality. Only two cited studies found an association between use of anticholinergic medications and reduced cognitive performance; however, these did not meet our inclusion criteria due to crude assessment of cognition and no adjustment for confounders.

### Bladder anticholinergic drug studies

#### Risk of dementia and cognitive outcomes

Fifteen studies investigating specific anticholinergic medications for bladder disorders in relation to dementia or cognitive outcomes are included in Supplementary Table 3. These varied in design including small randomised controlled trials (RCTs) such as Geller 2017^25^ (n=59), cohort studies such as Moga 2017^26^ (n=7,735) and large case-control studies using electronic health records such as Richardson 2018 (n=40,770 cases, 283,933 controls)^3^. All studies investigating risk of dementia in those taking any bladder drug (vs none) found a significant relationship; those examining scores on cognitive scales were more inconsistent in their findings.

Barthold 2020^27^ compared agents with selective antimuscarinic action at the M_3_ receptor, (solifenacin and darifenacin) with non-selective agents (oxybutynin, tolterodine, trospium, fesoterodine), but found no difference in dementia risk. Malcher 2022^28^ found that OAB anticholinergic use was associated with an increased risk of dementia with a cumulative dose response and when comparing drugs, they found a particularly marked increased risk of dementia for oxybutynin and solifenacin, but no increased risk for trospium. In contrast, Matta 2021^29^ found that receipt of solifenacin (OR 1.34), darifenacin (OR 1.49), tolterodine (OR 1.21), and fesoterodine (OR 1.39) were associated with increased odds of incident dementia compared with receipt of mirabegron. However, no effect was seen with oxybutynin or trospium.

Our search identified 13 reviews estimating the risks associated with anticholinergic agents used for Overactive Bladder (OAB). Most reviews were not systematic. One systematic review by Triantafylidis 2018^30^ investigated combined cholinesterase inhibitors with bladder anticholinergic drugs and found mixed results for cognitive and functional outcomes, another by Paquette 2011^31^ investigated reporting bias associated with adverse central nervous system events in clinical drug trials of younger and older adults with overactive bladder; however, study heterogeneity, dosing inconsistency, and reporting bias limited interpretation of the findings from the meta-analyses. Rangganata 2020^32^ conducted a systematic review and meta-analysis of reported effects on cognitive function in older people, finding that oxybutynin had the strongest association with MMSE decline, with weaker but statistically significant associations for darifenacin and tolterodine.

#### Mortality and hospitalisation in people with dementia

Four studies examining these relationships from the UK, US, and Northern Ireland (see Table 3) were included. McMichael 2021^33^ reported an association between urological drugs and increased mortality in people with dementia, but did not compare different bladder agents. Kachru 2020^34^ compared M_3_ selective (solifenacin, darifenacin) and the remaining non-selective bladder antimuscarinic agents and found a 50% higher risk of 180-day mortality associated with non-selective drugs, consistent across multiple sensitivity analyses. Similarly, Bishara 2021^35^ found that bladder drugs with a high central anticholinergic burden (tolterodine, oxybutynin) were associated with a 55% increased mortality risk compared to drugs with a low or zero score (darifenacin, fesoterodine, trospium, mirabegron, solifenacin).

**Table 3.**
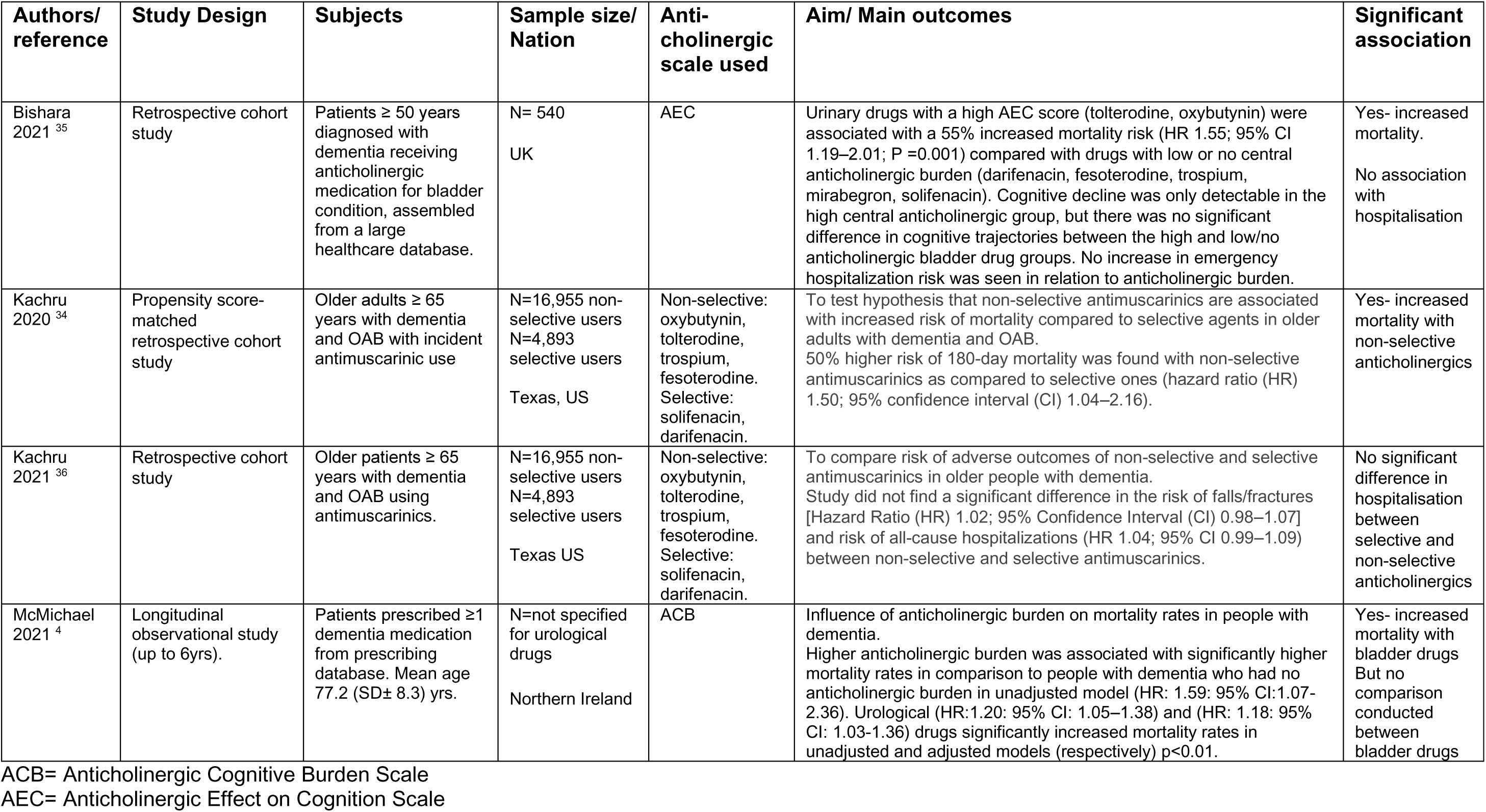
Papers examining the association between bladder anticholinergics and mortality or hospitalisation in people with dementia.

Kachru 2021^36^ found no increased risk of hospitalisation in patients receiving non-selective M_3_ medications compared to selective agents and Bishara^35^ found no excess hospitalisation in patients prescribed bladder anticholinergics with high central anticholinergic burden.

### Antidepressant investigations

#### Risk of dementia and cognitive outcomes

Supplementary Table 4 displays seven studies that specifically investigated anticholinergic antidepressants: mainly large cohort or population-based database studies. All four studies examining risk of dementia found an association between anticholinergic antidepressants and increased risk^2 3 37 38^. However, the three^39–41^ investigating cognition (whether in a specific population^40^ or in people with dementia^39^) found no association. No reviews were identified from our search.

#### Mortality or hospitalisation in dementia

Studies investigating the relationship between anticholinergic burden of antidepressants and outcomes in dementia are shown in Table 4. McMichael 2021^33^ found that antidepressant drugs were not significantly associated with mortality rates in people with dementia but did not compare the risk between different antidepressants or by anticholinergic burden. Mate 2022^40^ found no association with increased hospitalisation. Bishara 2021^39^ found a *reduced* risk of mortality in people receiving antidepressants with high central anticholinergic burden compared to those with no or low burden. No significant associations were detected between antidepressant’s central anticholinergic burden and hospitalisation (or cognitive decline).

**Table 4.**
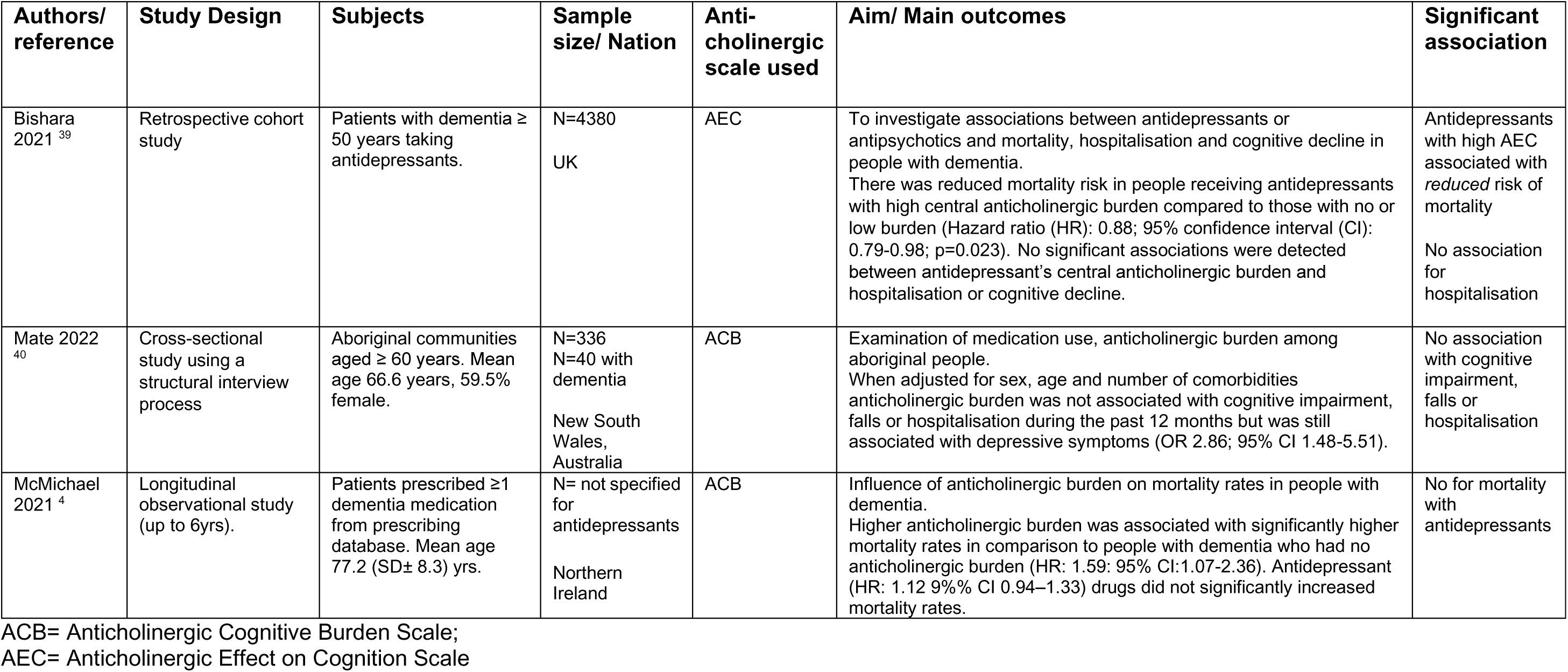
Papers examining the association between antidepressants with high anticholinergic burden and risk of mortality or hospitalisation in people with dementia.

## Discussion

Our review found consistent associations between certain classes of drugs with anticholinergic action (antidepressants and urological drugs) as well as anticholinergic burden and future dementia incidence^2 3^ underscoring the importance of anticholinergic drugs as a potential modifiable risk factor for dementia prevention^6^.

Evidence for an effect on cognitive outcomes in community populations was considerably less consistent. Regarding studies investigating outcomes in people with dementia, it appears that whilst significant associations have been reported between anticholinergic burden and both mortality and hospitalisation, no papers examined found an association between anticholinergic burden and longitudinal cognitive trajectories.

An association between anticholinergic burden and cognitive outcomes has not been conclusively shown for people with or without dementia. This review, as well as others^8 9^ highlight the complexities of assessing the long-term cognitive effects of anticholinergic drugs: measurement issues, reporting issues and confounding by indication. For instance, inconsistencies between studies may reflect the limited accuracy and consistency in the measurements used, or the way the results are reported. Even when using the same measure, different studies may report cognitive decline (change in cognitive function over time), cognitive impairment (cognitive function below a given level at a given time), transition to mild cognitive impairment (MCI), cognitive or neuropsychological performance etc. Furthermore, the measures themselves have limitations. MMSE scores are known to be limited by “floor effects” in those with more severe dementia and “ceiling effects” when used in individuals with a high educational background^42^. Also, recording of serial MMSE scores from clinical records can also be unreliable, which is an important limitation to the evaluations of cognitive function in studies. Cognitive decline measured by a change or cut-off based on cognitive tests, may have questionable clinical relevance as the tests may be insufficiently sensitive to detect cognitive decline among older adults^8^. In conclusion, a causal link between anticholinergic agents and cognitive decline cannot yet be inferred from these observational studies with considerable risk of bias, although the consistent finding of association with incident dementia may be enough for some clinicians to act on.

This review also examined papers investigating bladder drugs specifically and found again a consistent association between the use of anticholinergic agents used in OAB and the risk of dementia; however, the association with cognitive decline is also less clear. In addition to the reasons already discussed, this could also be because newer agents less likely to penetrate the BBB and to act centrally were used in most studies.

Supporting this, in studies investigating dementia outcomes, two studies compared bladder agents and found that non-selective agents or centrally acting drugs (oxybutynin, tolterodine) were associated with a 50%^34^ and 55%^35^ increased mortality risk than the more selective agents or those with lower central activity. However, there was no association within this group with high central anticholinergic burden and hospitalisation. This may be due to the non-differential effects of the muscarinic receptors on falls/fractures and all-cause hospitalisation^36^. Another reason might be that muscarinic selectivity is not binary, meaning the drugs categorised as selective to M_3_ receptors still have some activity with the M_1_ receptors, albeit weaker than the non-selective drugs, and that this activity may still be strong enough to be harmful^27^.

With regards to the papers exploring the relationships with antidepressants, whilst anticholinergic antidepressants were associated with increased risk of dementia in some studies, they were also not associated with cognitive decline, although this could reflect the treatment of depression resulting in improved cognition, even when anticholinergic agents are used^43^. In addition, antidepressant drugs with high anticholinergic burden were not significantly associated with increased mortality rates in people with dementia^4^; if anything, risks were reduced^39^. Furthermore, no significant associations were detected between antidepressant’s central anticholinergic burden and hospitalisation (or cognitive decline)^39 40^. While further data are required, it appears that these counter-intuitive findings may reflect factors underlying the prescriber choice of antidepressants rather than the agents themselves^39^.

### Strengths and limitations

Strengths of this review include the wider inclusion of studies investigating anticholinergic medication and dementia outcomes alongside those investigating dementia risk, meaning that the evidence from both fields can be considered in combination to build up a better understanding of the overall risks involved. It is even more necessary to triangulate results with observational data.

Limitations include our use of only PubMed and Cochrane databases, although front and back referencing was carried out and references from all papers and reviews were scanned carefully so as not to miss any relevant studies. Equally, while we assessed paper quality when looking at the effect of anticholinergic medication on people with dementia, we did not extend this to other aspects of the review where there was sufficient existing review evidence.

Issues of quality with the evidence base are of note. The methods for collecting medication use, whilst more reliable with the use of electronic pharmacy dispensing data, may still not accurately reflect actual or continued use. Second, multiple risk scales are available for estimating anticholinergic burden of drugs and therefore studies cannot easily be compared nor combined.

It is unclear why there is a consistent association between anticholinergic burden and dementia risk however a similar relationship was not found with cognitive impairment/decline either in people with or without dementia. Higher dementia incidence may be occurring through a reduced ability to compensate in the early stages of neurodegeneration. Anticholinergic drugs could potentially bring forward the onset of dementia, by compromising brain function at borderline impairment, rather than directly influencing the underlying disease processes. This may therefore not manifest as detectable cognitive deficits. Moreover, mortality and hospitalisation outcomes in dementia might possibly reflect non-cognitive adverse consequences of anticholinergic actions. Urinary drugs with high anticholinergic burden including oxybutynin and tolterodine should be avoided in people with dementia due to the associated increased mortality risk identified in studies. Furthermore, whilst anticholinergic antidepressants are associated with increased risk of dementia, they were not found to increase mortality in dementia. Antidepressant studies are likely to be most subject to confounding by indication, which may explain these results; clinicians may avoid the more anticholinergic ones in people who are frail, have various comorbidities and who are already at higher risk of mortality whereas risks with urinary drugs may be less widely known. Further studies that can dissect the level at which anticholinergic agents act would be useful and could help better inform guides for safe usage in the future.

## Conclusion

Consistent associations were therefore found between anticholinergic burden and future dementia incidence. In people with dementia, anticholinergic burden was associated with increased mortality and hospitalisation. Associations with cognitive outcomes in people without/with dementia remains uncertain. Current evidence remains derived from observation studies and will need reconsideration as improved methodology is developed. However, it remains reasonable for clinicians to exercise caution with anticholinergic medication and carefully weigh up risks and benefits when making prescribing decisions.

## Data Availability

All data produced in the present work are contained in the manuscript

---

**Supplementary Table 1.**
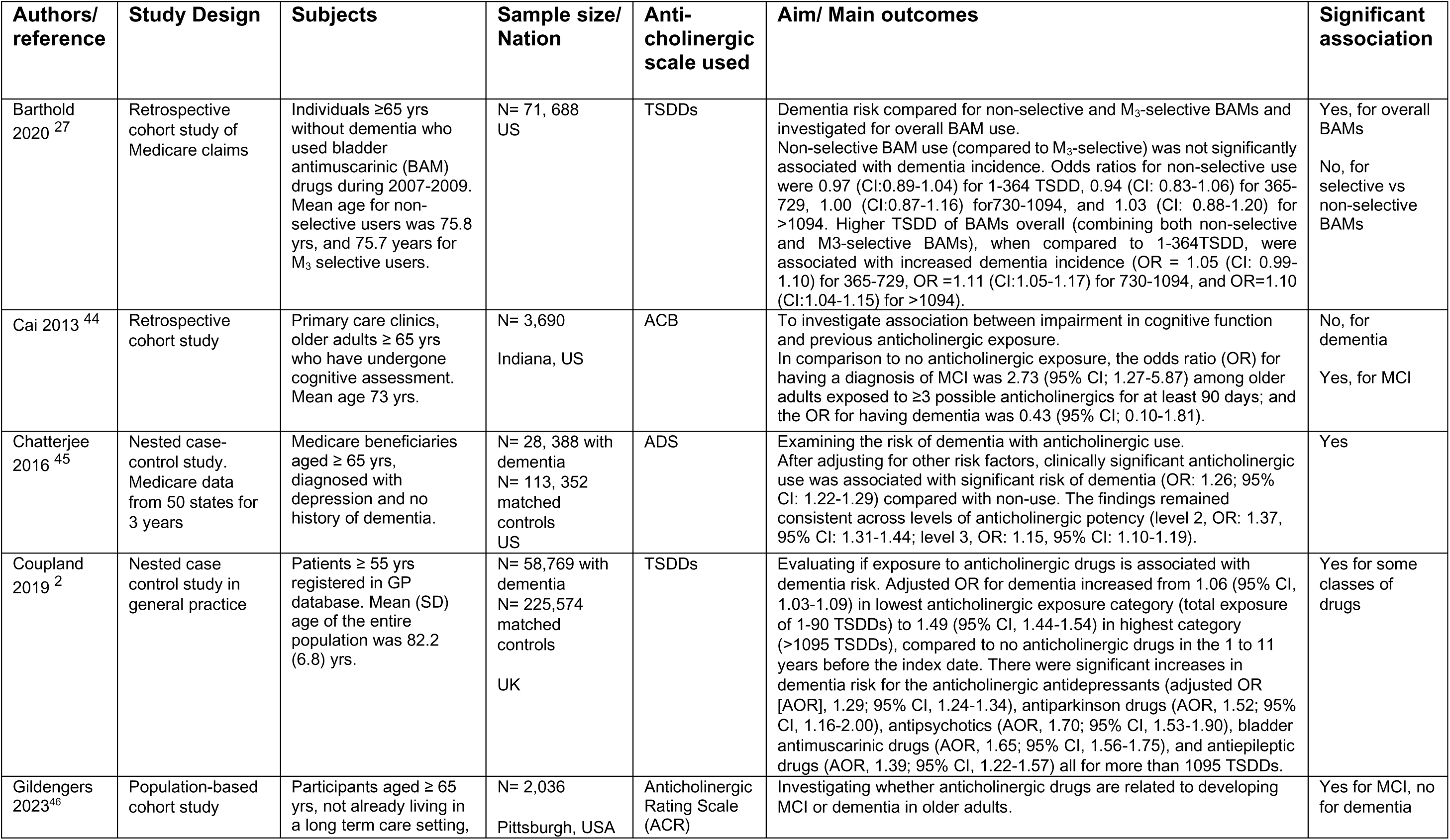
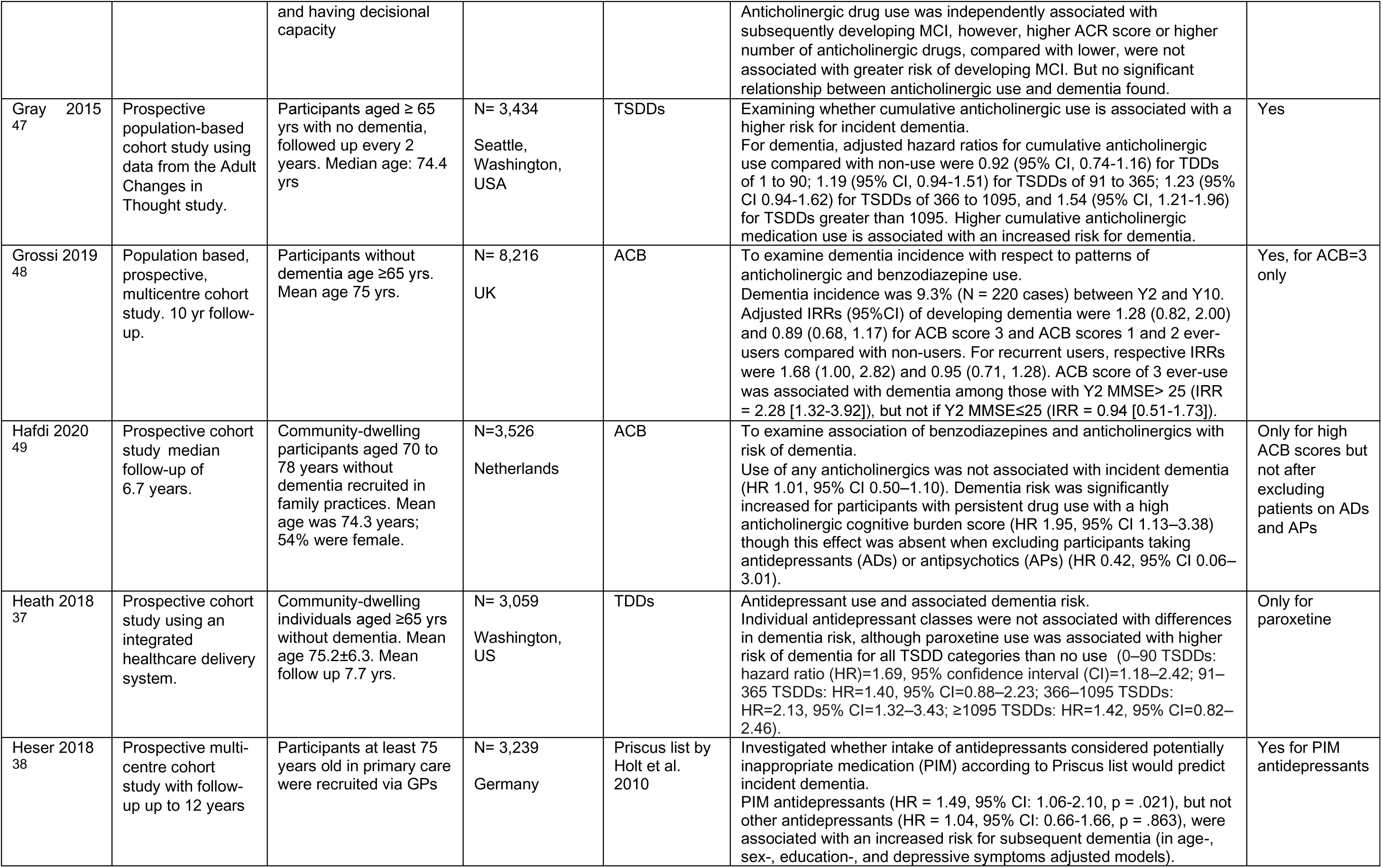
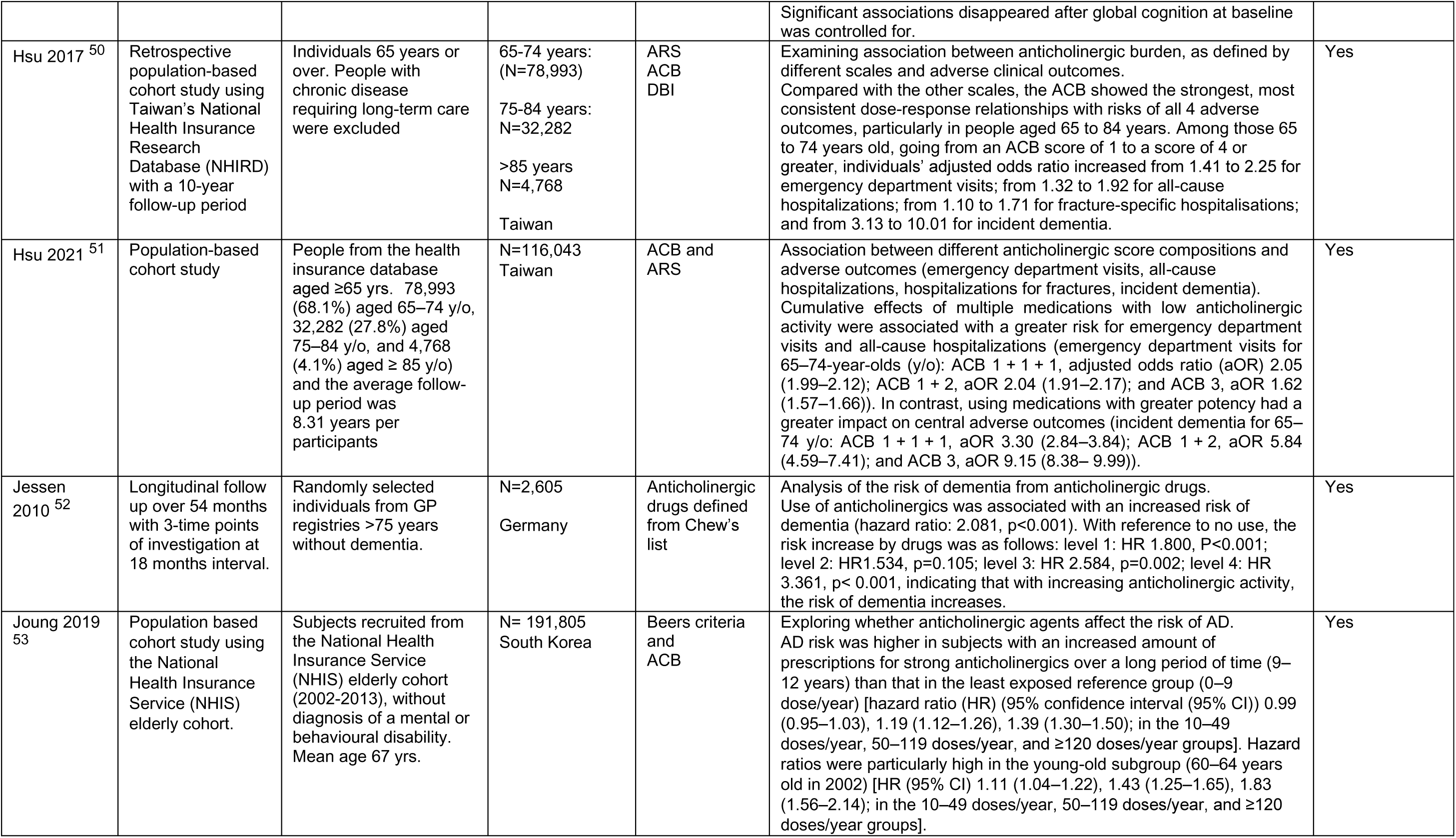
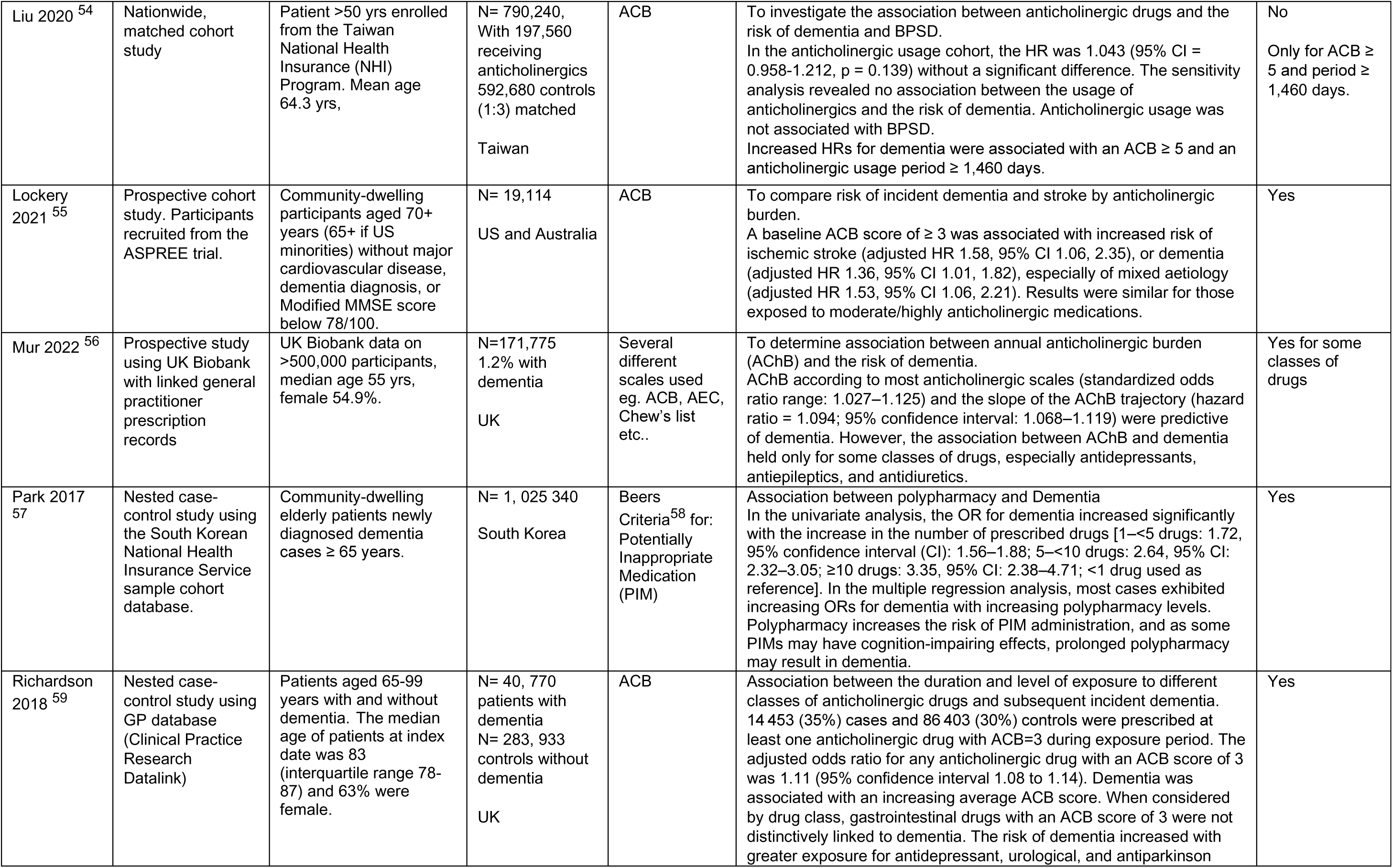
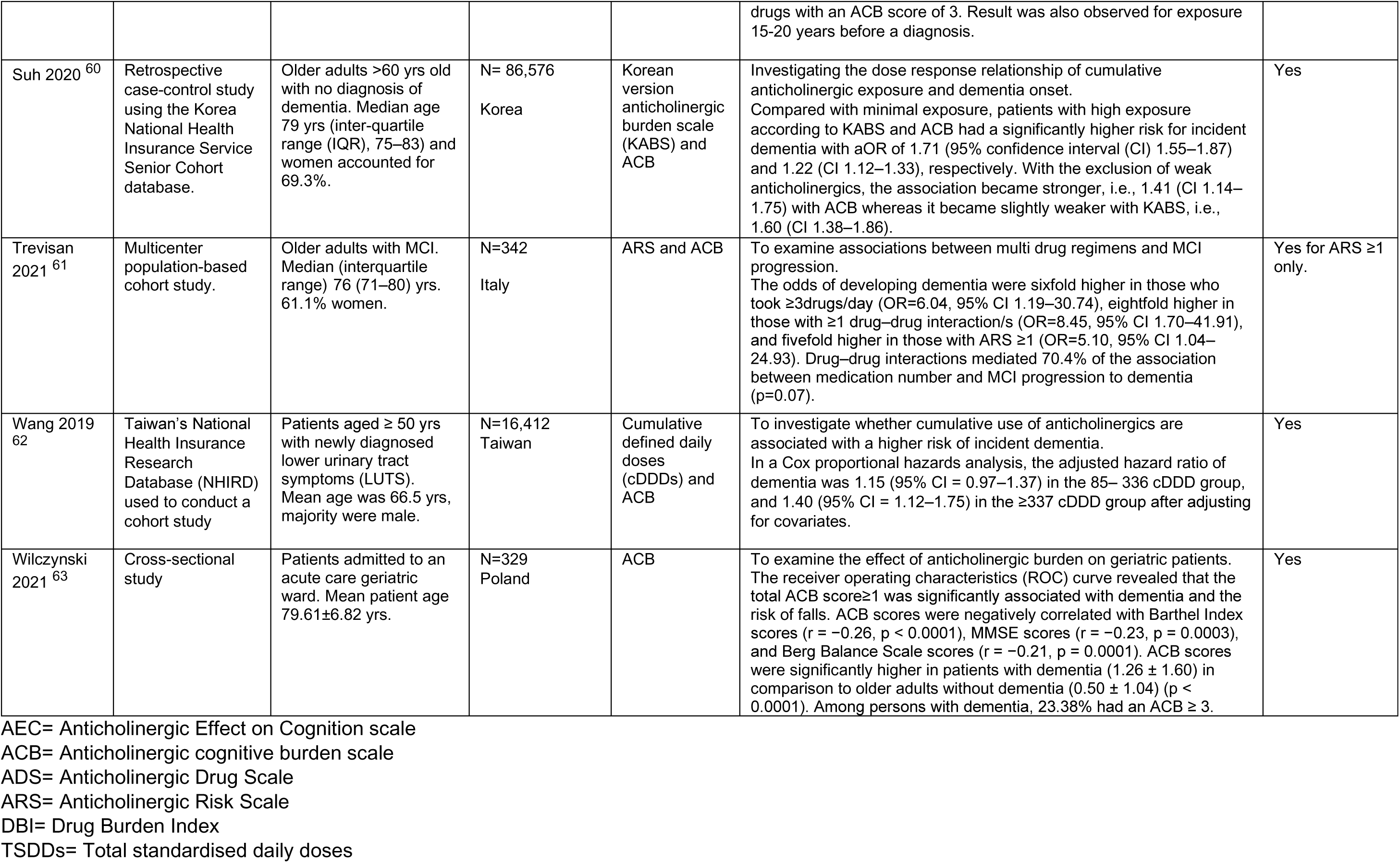
Papers examining the association between anticholinergic drugs and risk of dementia.

| Authors/<br>reference | Study Design | Subjects | Sample size/<br>Nation | Anti-<br>cholinergic<br>scale used | Aim/ Main outcomes | Significant<br>association |
| --- | --- | --- | --- | --- | --- | --- |
| Barthold<br>2020 <sup>27</sup> | Retrospective<br>cohort study of<br>Medicare claims | Individuals ≥65 yrs<br>without dementia who<br>used bladder<br>antimuscarinic (BAM)<br>drugs during 2007-2009.<br>Mean age for non-selective users was 75.8<br>yrs, and 75.7 years for<br>M<sub>3</sub> selective users. | N= 71, 688<br>US | TSDDs | Dementia risk compared for non-selective and M<sub>3</sub>-selective BAMs and investigated for overall BAM use.<br>Non-selective BAM use (compared to M<sub>3</sub>-selective) was not significantly associated with dementia incidence. Odds ratios for non-selective use were 0.97 (CI:0.89-1.04) for 1-364 TSDD, 0.94 (CI: 0.83-1.06) for 365-729, 1.00 (CI:0.87-1.16) for 730-1094, and 1.03 (CI: 0.88-1.20) for >1094. Higher TSDD of BAMs overall (combining both non-selective and M<sub>3</sub>-selective BAMs), when compared to 1-364TSDD, were associated with increased dementia incidence (OR = 1.05 (CI: 0.99-1.10) for 365-729, OR =1.11 (CI:1.05-1.17) for 730-1094, and OR=1.10 (CI:1.04-1.15) for >1094). | Yes, for overall<br>BAMs<br>No, for<br>selective vs<br>non-selective<br>BAMs |
| Cai 2013 <sup>44</sup> | Retrospective<br>cohort study | Primary care clinics,<br>older adults ≥ 65 yrs<br>who have undergone<br>cognitive assessment.<br>Mean age 73 yrs. | N= 3,690<br>Indiana, US | ACB | To investigate association between impairment in cognitive function and previous anticholinergic exposure.<br>In comparison to no anticholinergic exposure, the odds ratio (OR) for having a diagnosis of MCI was 2.73 (95% CI; 1.27-5.87) among older adults exposed to ≥3 possible anticholinergics for at least 90 days; and the OR for having dementia was 0.43 (95% CI; 0.10-1.81). | No, for<br>dementia<br>Yes, for MCI |
| Chatterjee<br>2016 <sup>45</sup> | Nested case-control study.<br>Medicare data<br>from 50 states for<br>3 years | Medicare beneficiaries<br>aged ≥ 65 yrs,<br>diagnosed with<br>depression and no<br>history of dementia. | N= 28, 388 with<br>dementia<br>N= 113, 352<br>matched<br>controls<br>US | ADS | Examining the risk of dementia with anticholinergic use.<br>After adjusting for other risk factors, clinically significant anticholinergic use was associated with significant risk of dementia (OR: 1.26; 95% CI: 1.22-1.29) compared with non-use. The findings remained consistent across levels of anticholinergic potency (level 2, OR: 1.37, 95% CI: 1.31-1.44; level 3, OR: 1.15, 95% CI: 1.10-1.19). | Yes |
| Coupland<br>2019 <sup>2</sup> | Nested case<br>control study in<br>general practice | Patients ≥ 55 yrs<br>registered in GP<br>database. Mean (SD)<br>age of the entire<br>population was 82.2<br>(6.8) yrs. | N= 58,769 with<br>dementia<br>N= 225,574<br>matched<br>controls<br>UK | TSDDs | Evaluating if exposure to anticholinergic drugs is associated with dementia risk. Adjusted OR for dementia increased from 1.06 (95% CI, 1.03-1.09) in lowest anticholinergic exposure category (total exposure of 1-90 TSDDs) to 1.49 (95% CI, 1.44-1.54) in highest category (>1095 TSDDs), compared to no anticholinergic drugs in the 1 to 11 years before the index date. There were significant increases in dementia risk for the anticholinergic antidepressants (adjusted OR [AOR], 1.29; 95% CI, 1.24-1.34), antiparkinson drugs (AOR, 1.52; 95% CI, 1.16-2.00), antipsychotics (AOR, 1.70; 95% CI, 1.53-1.90), bladder antimuscarinic drugs (AOR, 1.65; 95% CI, 1.56-1.75), and antiepileptic drugs (AOR, 1.39; 95% CI, 1.22-1.57) all for more than 1095 TSDDs. | Yes for some<br>classes of<br>drugs |
| Gildengers<br>2023<sup>46</sup> | Population-based<br>cohort study | Participants aged ≥ 65<br>yrs, not already living in<br>a long term care setting, | N= 2,036<br>Pittsburgh, USA | Anticholinergic<br>Rating Scale<br>(ACR) | Investigating whether anticholinergic drugs are related to developing MCI or dementia in older adults. | Yes for MCI, no<br>for dementia |
|  |  | and having decisional capacity |  |  | Anticholinergic drug use was independently associated with subsequently developing MCI, however, higher ACR score or higher number of anticholinergic drugs, compared with lower, were not associated with greater risk of developing MCI. But no significant relationship between anticholinergic use and dementia found. |  |
| Gray 2015<br>47 | Prospective population-based cohort study using data from the Adult Changes in Thought study. | Participants aged ≥ 65 yrs with no dementia, followed up every 2 years. Median age: 74.4 yrs | N= 3,434<br>Seattle, Washington, USA | TSDDs | Examining whether cumulative anticholinergic use is associated with a higher risk for incident dementia.<br>For dementia, adjusted hazard ratios for cumulative anticholinergic use compared with non-use were 0.92 (95% CI, 0.74-1.16) for TDDs of 1 to 90; 1.19 (95% CI, 0.94-1.51) for TSDDs of 91 to 365; 1.23 (95% CI 0.94-1.62) for TSDDs of 366 to 1095, and 1.54 (95% CI, 1.21-1.96) for TSDDs greater than 1095. Higher cumulative anticholinergic medication use is associated with an increased risk for dementia. | Yes |
| Grossi 2019<br>48 | Population based, prospective, multicentre cohort study. 10 yr follow-up. | Participants without dementia age ≥65 yrs. Mean age 75 yrs. | N= 8,216<br>UK | ACB | To examine dementia incidence with respect to patterns of anticholinergic and benzodiazepine use.<br>Dementia incidence was 9.3% (N = 220 cases) between Y2 and Y10. Adjusted IRRs (95%CI) of developing dementia were 1.28 (0.82, 2.00) and 0.89 (0.68, 1.17) for ACB score 3 and ACB scores 1 and 2 ever-users compared with non-users. For recurrent users, respective IRRs were 1.68 (1.00, 2.82) and 0.95 (0.71, 1.28). ACB score of 3 ever-use was associated with dementia among those with Y2 MMSE> 25 (IRR = 2.28 [1.32-3.92]), but not if Y2 MMSE≤25 (IRR = 0.94 [0.51-1.73]). | Yes, for ACB=3 only |
| Hafdi 2020<br>49 | Prospective cohort study median follow-up of 6.7 years. | Community-dwelling participants aged 70 to 78 years without dementia recruited in family practices. Mean age was 74.3 years; 54% were female. | N=3,526<br>Netherlands | ACB | To examine association of benzodiazepines and anticholinergics with risk of dementia.<br>Use of any anticholinergics was not associated with incident dementia (HR 1.01, 95% CI 0.50–1.10). Dementia risk was significantly increased for participants with persistent drug use with a high anticholinergic cognitive burden score (HR 1.95, 95% CI 1.13–3.38) though this effect was absent when excluding participants taking antidepressants (ADs) or antipsychotics (APs) (HR 0.42, 95% CI 0.06–3.01). | Only for high ACB scores but not after excluding patients on ADs and APs |
| Heath 2018<br>37 | Prospective cohort study using an integrated healthcare delivery system. | Community-dwelling individuals aged ≥65 yrs without dementia. Mean age 75.2±6.3. Mean follow up 7.7 yrs. | N= 3,059<br>Washington, US | TDDs | Antidepressant use and associated dementia risk.<br>Individual antidepressant classes were not associated with differences in dementia risk, although paroxetine use was associated with higher risk of dementia for all TSDD categories than no use (0–90 TSDDs: hazard ratio (HR)=1.69, 95% confidence interval (CI)=1.18–2.42; 91–365 TSDDs: HR=1.40, 95% CI=0.88–2.23; 366–1095 TSDDs: HR=2.13, 95% CI=1.32–3.43; ≥1095 TSDDs: HR=1.42, 95% CI=0.82–2.46). | Only for paroxetine |
| Heser 2018<br>38 | Prospective multi-centre cohort study with follow-up up to 12 years | Participants at least 75 years old in primary care were recruited via GPs | N= 3,239<br>Germany | Priscus list by Holt et al. 2010 | Investigated whether intake of antidepressants considered potentially inappropriate medication (PIM) according to Priscus list would predict incident dementia.<br>PIM antidepressants (HR = 1.49, 95% CI: 1.06-2.10, p = .021), but not other antidepressants (HR = 1.04, 95% CI: 0.66-1.66, p = .863), were associated with an increased risk for subsequent dementia (in age-, sex-, education-, and depressive symptoms adjusted models). | Yes for PIM antidepressants |
|  |  |  |  |  | Significant associations disappeared after global cognition at baseline was controlled for. |  |
| Hsu 2017 <sup>50</sup> | Retrospective population-based cohort study using Taiwan's National Health Insurance Research Database (NHIRD) with a 10-year follow-up period | Individuals 65 years or over. People with chronic disease requiring long-term care were excluded | 65-74 years: (N=78,993)<br>75-84 years: N=32,282<br>>85 years N=4,768<br>Taiwan | ARS<br>ACB<br>DBI | Examining association between anticholinergic burden, as defined by different scales and adverse clinical outcomes. Compared with the other scales, the ACB showed the strongest, most consistent dose-response relationships with risks of all 4 adverse outcomes, particularly in people aged 65 to 84 years. Among those 65 to 74 years old, going from an ACB score of 1 to a score of 4 or greater, individuals' adjusted odds ratio increased from 1.41 to 2.25 for emergency department visits; from 1.32 to 1.92 for all-cause hospitalizations; from 1.10 to 1.71 for fracture-specific hospitalisations; and from 3.13 to 10.01 for incident dementia. | Yes |
| Hsu 2021 <sup>51</sup> | Population-based cohort study | People from the health insurance database aged ≥65 yrs. 78,993 (68.1%) aged 65–74 y/o, 32,282 (27.8%) aged 75–84 y/o, and 4,768 (4.1%) aged ≥ 85 y/o) and the average follow-up period was 8.31 years per participants | N=116,043<br>Taiwan | ACB and<br>ARS | Association between different anticholinergic score compositions and adverse outcomes (emergency department visits, all-cause hospitalizations, hospitalizations for fractures, incident dementia). Cumulative effects of multiple medications with low anticholinergic activity were associated with a greater risk for emergency department visits and all-cause hospitalizations (emergency department visits for 65–74-year-olds (y/o): ACB 1 + 1 + 1, adjusted odds ratio (aOR) 2.05 (1.99–2.12); ACB 1 + 2, aOR 2.04 (1.91–2.17); and ACB 3, aOR 1.62 (1.57–1.66)). In contrast, using medications with greater potency had a greater impact on central adverse outcomes (incident dementia for 65–74 y/o: ACB 1 + 1 + 1, aOR 3.30 (2.84–3.84); ACB 1 + 2, aOR 5.84 (4.59–7.41); and ACB 3, aOR 9.15 (8.38– 9.99)). | Yes |
| Jessen 2010 <sup>52</sup> | Longitudinal follow up over 54 months with 3-time points of investigation at 18 months interval. | Randomly selected individuals from GP registries >75 years without dementia. | N=2,605<br>Germany | Anticholinergic drugs defined from Chew's list | Analysis of the risk of dementia from anticholinergic drugs. Use of anticholinergics was associated with an increased risk of dementia (hazard ratio: 2.081, p<0.001). With reference to no use, the risk increase by drugs was as follows: level 1: HR 1.800, P<0.001; level 2: HR1.534, p=0.105; level 3: HR 2.584, p=0.002; level 4: HR 3.361, p< 0.001, indicating that with increasing anticholinergic activity, the risk of dementia increases. | Yes |
| Joung 2019 <sup>53</sup> | Population based cohort study using the National Health Insurance Service (NHIS) elderly cohort. | Subjects recruited from the National Health Insurance Service (NHIS) elderly cohort (2002-2013), without diagnosis of a mental or behavioural disability. Mean age 67 yrs. | N= 191,805<br>South Korea | Beers criteria and<br>ACB | Exploring whether anticholinergic agents affect the risk of AD. AD risk was higher in subjects with an increased amount of prescriptions for strong anticholinergics over a long period of time (9–12 years) than that in the least exposed reference group (0–9 dose/year) [hazard ratio (HR) (95% confidence interval (95% CI)) 0.99 (0.95–1.03), 1.19 (1.12–1.26), 1.39 (1.30–1.50); in the 10–49 doses/year, 50–119 doses/year, and ≥120 doses/year groups]. Hazard ratios were particularly high in the young-old subgroup (60–64 years old in 2002) [HR (95% CI) 1.11 (1.04–1.22), 1.43 (1.25–1.65), 1.83 (1.56–2.14); in the 10–49 doses/year, 50–119 doses/year, and ≥120 doses/year groups]. | Yes |
| Liu 2020<sup>54</sup> | Nationwide, matched cohort study | Patient >50 yrs enrolled from the Taiwan National Health Insurance (NHI) Program. Mean age 64.3 yrs, | N= 790,240, With 197,560 receiving anticholinergics 592,680 controls (1:3) matched<br>Taiwan | ACB | To investigate the association between anticholinergic drugs and the risk of dementia and BPSD.<br>In the anticholinergic usage cohort, the HR was 1.043 (95% CI = 0.958-1.212, p = 0.139) without a significant difference. The sensitivity analysis revealed no association between the usage of anticholinergics and the risk of dementia. Anticholinergic usage was not associated with BPSD.<br>Increased HRs for dementia were associated with an ACB ≥ 5 and an anticholinergic usage period ≥ 1,460 days. | No<br>Only for ACB ≥ 5 and period ≥ 1,460 days. |
| Lockery 2021<sup>55</sup> | Prospective cohort study. Participants recruited from the ASPREE trial. | Community-dwelling participants aged 70+ years (65+ if US minorities) without major cardiovascular disease, dementia diagnosis, or Modified MMSE score below 78/100. | N= 19,114<br>US and Australia | ACB | To compare risk of incident dementia and stroke by anticholinergic burden.<br>A baseline ACB score of ≥ 3 was associated with increased risk of ischemic stroke (adjusted HR 1.58, 95% CI 1.06, 2.35), or dementia (adjusted HR 1.36, 95% CI 1.01, 1.82), especially of mixed aetiology (adjusted HR 1.53, 95% CI 1.06, 2.21). Results were similar for those exposed to moderate/highly anticholinergic medications. | Yes |
| Mur 2022<sup>56</sup> | Prospective study using UK Biobank with linked general practitioner prescription records | UK Biobank data on >500,000 participants, median age 55 yrs, female 54.9%. | N=171,775<br>1.2% with dementia<br>UK | Several different scales used eg. ACB, AEC, Chew's list etc.. | To determine association between annual anticholinergic burden (AChB) and the risk of dementia.<br>AChB according to most anticholinergic scales (standardized odds ratio range: 1.027–1.125) and the slope of the AChB trajectory (hazard ratio = 1.094; 95% confidence interval: 1.068–1.119) were predictive of dementia. However, the association between AChB and dementia held only for some classes of drugs, especially antidepressants, antiepileptics, and antidiuretics. | Yes for some classes of drugs |
| Park 2017<sup>57</sup> | Nested case-control study using the South Korean National Health Insurance Service sample cohort database. | Community-dwelling elderly patients newly diagnosed dementia cases ≥ 65 years. | N= 1, 025 340<br>South Korea | Beers Criteria<sup>58</sup> for: Potentially Inappropriate Medication (PIM) | Association between polypharmacy and Dementia<br>In the univariate analysis, the OR for dementia increased significantly with the increase in the number of prescribed drugs [1–<5 drugs: 1.72, 95% confidence interval (CI): 1.56–1.88; 5–<10 drugs: 2.64, 95% CI: 2.32–3.05; ≥10 drugs: 3.35, 95% CI: 2.38–4.71; <1 drug used as reference]. In the multiple regression analysis, most cases exhibited increasing ORs for dementia with increasing polypharmacy levels. Polypharmacy increases the risk of PIM administration, and as some PIMs may have cognition-impairing effects, prolonged polypharmacy may result in dementia. | Yes |
| Richardson 2018<sup>59</sup> | Nested case-control study using GP database (Clinical Practice Research Datalink) | Patients aged 65-99 years with and without dementia. The median age of patients at index date was 83 (interquartile range 78-87) and 63% were female. | N= 40, 770 patients with dementia<br>N= 283, 933 controls without dementia<br>UK | ACB | Association between the duration and level of exposure to different classes of anticholinergic drugs and subsequent incident dementia.<br>14 453 (35%) cases and 86 403 (30%) controls were prescribed at least one anticholinergic drug with ACB=3 during exposure period. The adjusted odds ratio for any anticholinergic drug with an ACB score of 3 was 1.11 (95% confidence interval 1.08 to 1.14). Dementia was associated with an increasing average ACB score. When considered by drug class, gastrointestinal drugs with an ACB score of 3 were not distinctively linked to dementia. The risk of dementia increased with greater exposure for antidepressant, urological, and antiparkinson drugs with an ACB score of 3. Result was also observed for exposure 15-20 years before a diagnosis. | Yes |
| Suh 2020 <sup>60</sup> | Retrospective case-control study using the Korea National Health Insurance Service Senior Cohort database. | Older adults >60 yrs old with no diagnosis of dementia. Median age 79 yrs (inter-quartile range (IQR), 75–83) and women accounted for 69.3%. | N= 86,576<br>Korea | Korean version anticholinergic burden scale (KABS) and ACB | Investigating the dose response relationship of cumulative anticholinergic exposure and dementia onset. Compared with minimal exposure, patients with high exposure according to KABS and ACB had a significantly higher risk for incident dementia with aOR of 1.71 (95% confidence interval (CI) 1.55–1.87) and 1.22 (CI 1.12–1.33), respectively. With the exclusion of weak anticholinergics, the association became stronger, i.e., 1.41 (CI 1.14–1.75) with ACB whereas it became slightly weaker with KABS, i.e., 1.60 (CI 1.38–1.86). | Yes |
| Trevisan 2021 <sup>61</sup> | Multicenter population-based cohort study. | Older adults with MCI. Median (interquartile range) 76 (71–80) yrs. 61.1% women. | N=342<br>Italy | ARS and ACB | To examine associations between multi drug regimens and MCI progression. The odds of developing dementia were sixfold higher in those who took $\geq 3$ drugs/day (OR=6.04, 95% CI 1.19–30.74), eightfold higher in those with $\geq 1$ drug–drug interaction/s (OR=8.45, 95% CI 1.70–41.91), and fivefold higher in those with ARS $\geq 1$ (OR=5.10, 95% CI 1.04–24.93). Drug–drug interactions mediated 70.4% of the association between medication number and MCI progression to dementia (p=0.07). | Yes for ARS $\geq 1$ only. |
| Wang 2019 <sup>62</sup> | Taiwan's National Health Insurance Research Database (NHIRD) used to conduct a cohort study | Patients aged $\geq 50$ yrs with newly diagnosed lower urinary tract symptoms (LUTS). Mean age was 66.5 yrs, majority were male. | N=16,412<br>Taiwan | Cumulative defined daily doses (cDDD) and ACB | To investigate whether cumulative use of anticholinergics are associated with a higher risk of incident dementia. In a Cox proportional hazards analysis, the adjusted hazard ratio of dementia was 1.15 (95% CI = 0.97–1.37) in the 85– 336 cDDD group, and 1.40 (95% CI = 1.12–1.75) in the $\geq 337$ cDDD group after adjusting for covariates. | Yes |
| Wilczynski 2021 <sup>63</sup> | Cross-sectional study | Patients admitted to an acute care geriatric ward. Mean patient age 79.61±6.82 yrs. | N=329<br>Poland | ACB | To examine the effect of anticholinergic burden on geriatric patients. The receiver operating characteristics (ROC) curve revealed that the total ACB score $\geq 1$ was significantly associated with dementia and the risk of falls. ACB scores were negatively correlated with Barthel Index scores (r = -0.26, p < 0.0001), MMSE scores (r = -0.23, p = 0.0003), and Berg Balance Scale scores (r = -0.21, p = 0.0001). ACB scores were significantly higher in patients with dementia (1.26 ± 1.60) in comparison to older adults without dementia (0.50 ± 1.04) (p < 0.0001). Among persons with dementia, 23.38% had an ACB $\geq 3$. | Yes |

AEC= Anticholinergic Effect on Cognition scale
ACB= Anticholinergic cognitive burden scale
ADS= Anticholinergic Drug Scale
ARS= Anticholinergic Risk Scale
DBI= Drug Burden Index
TSDDs= Total standardised daily doses

**Supplementary Table 2.**
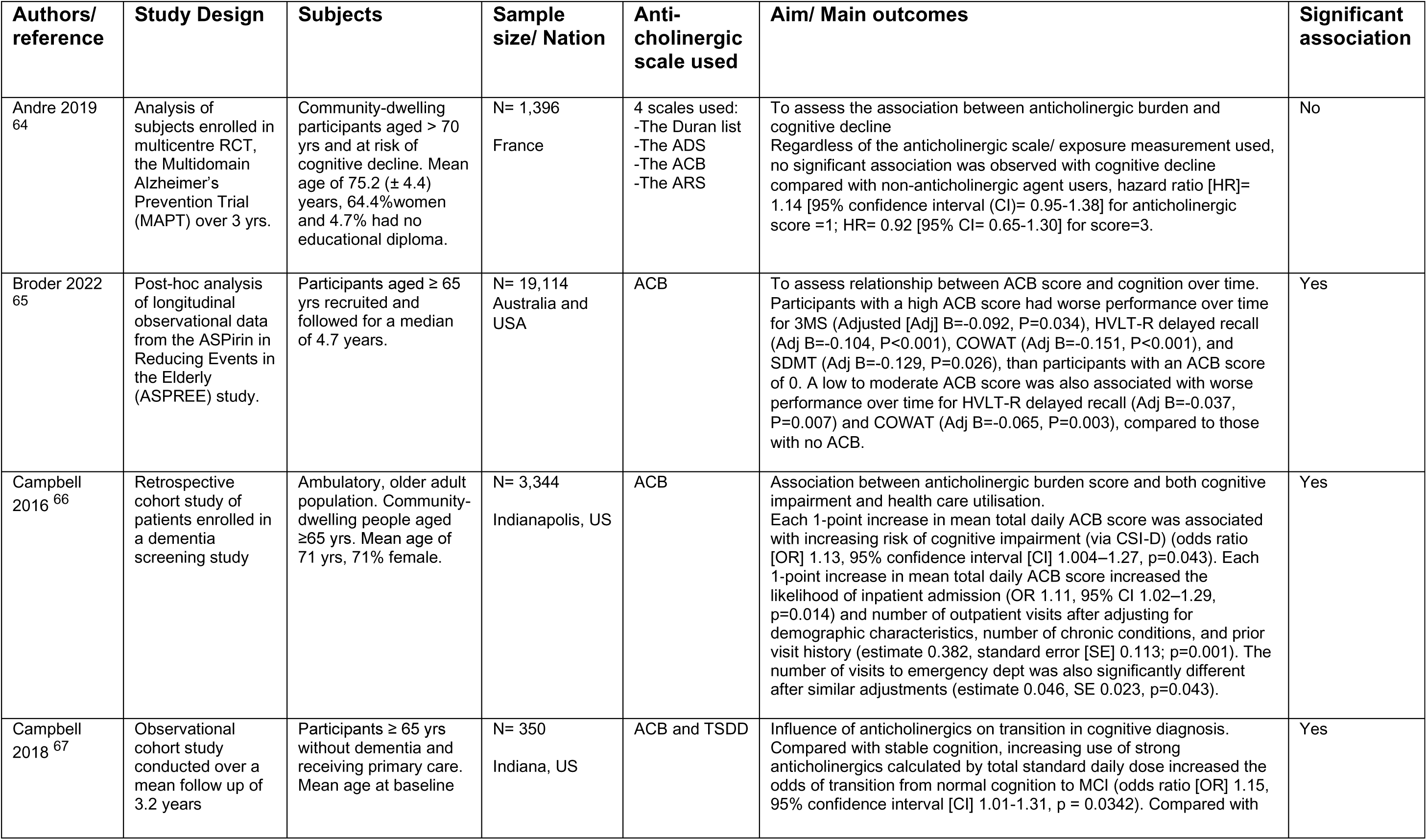
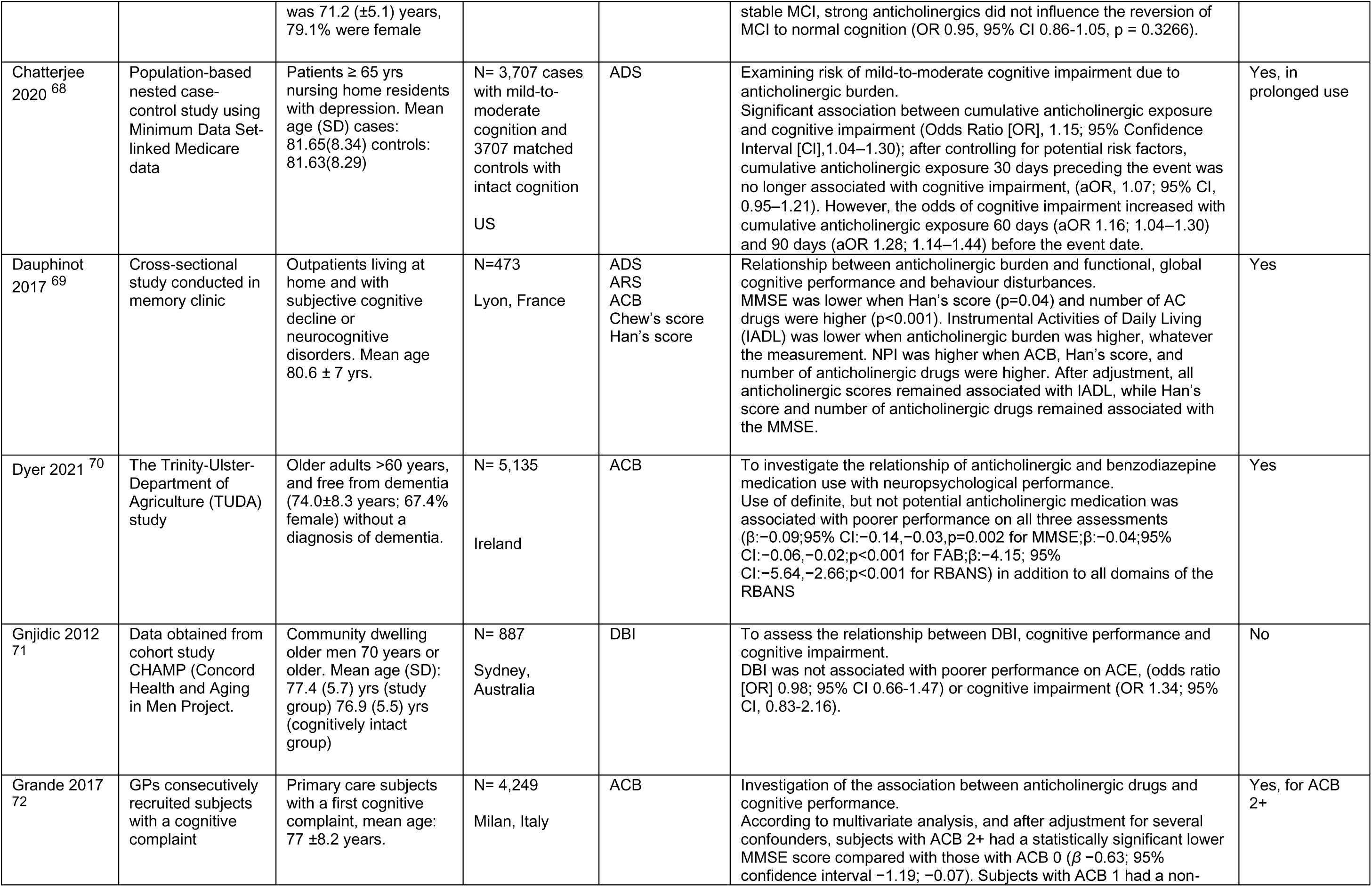
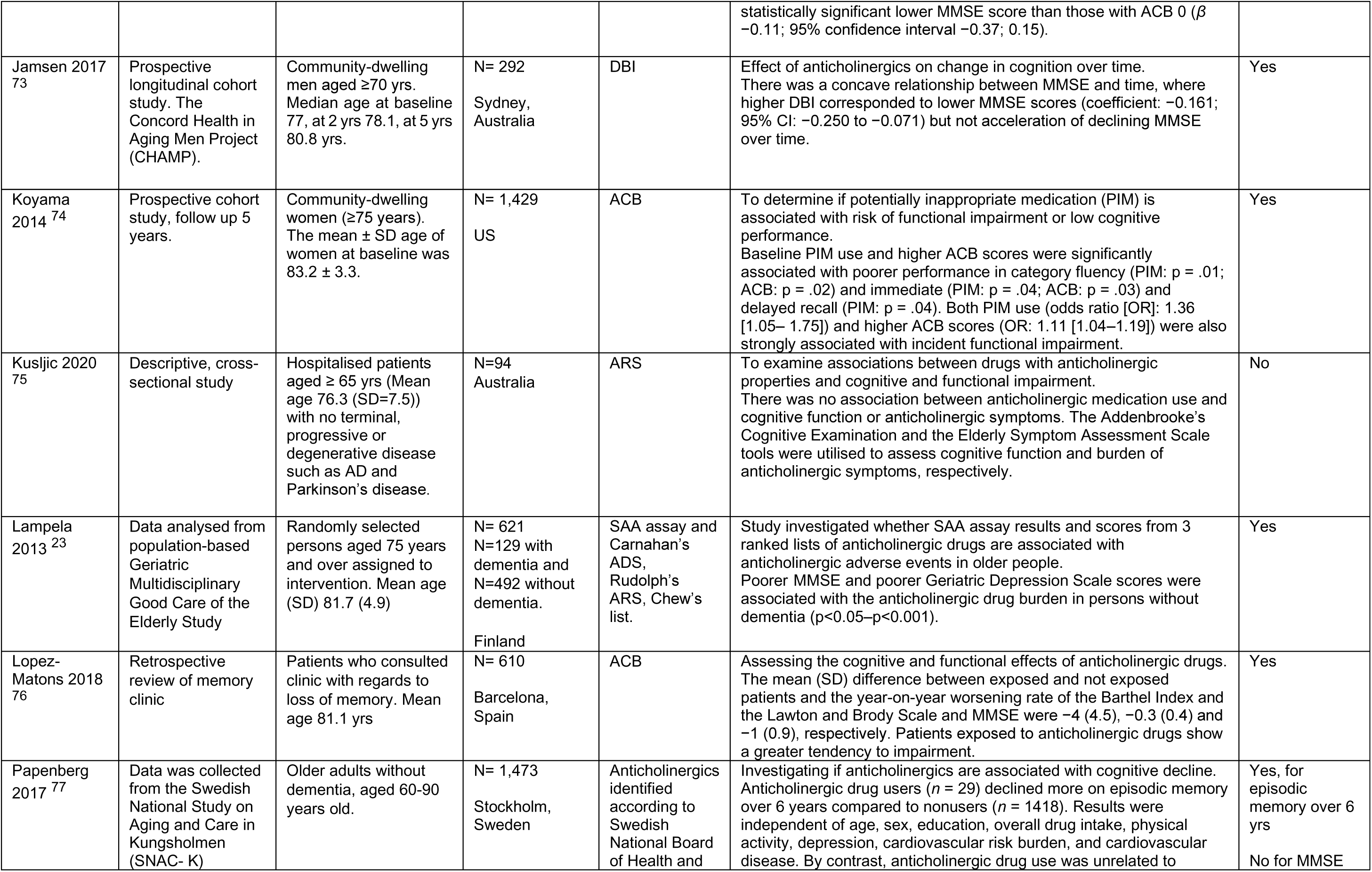
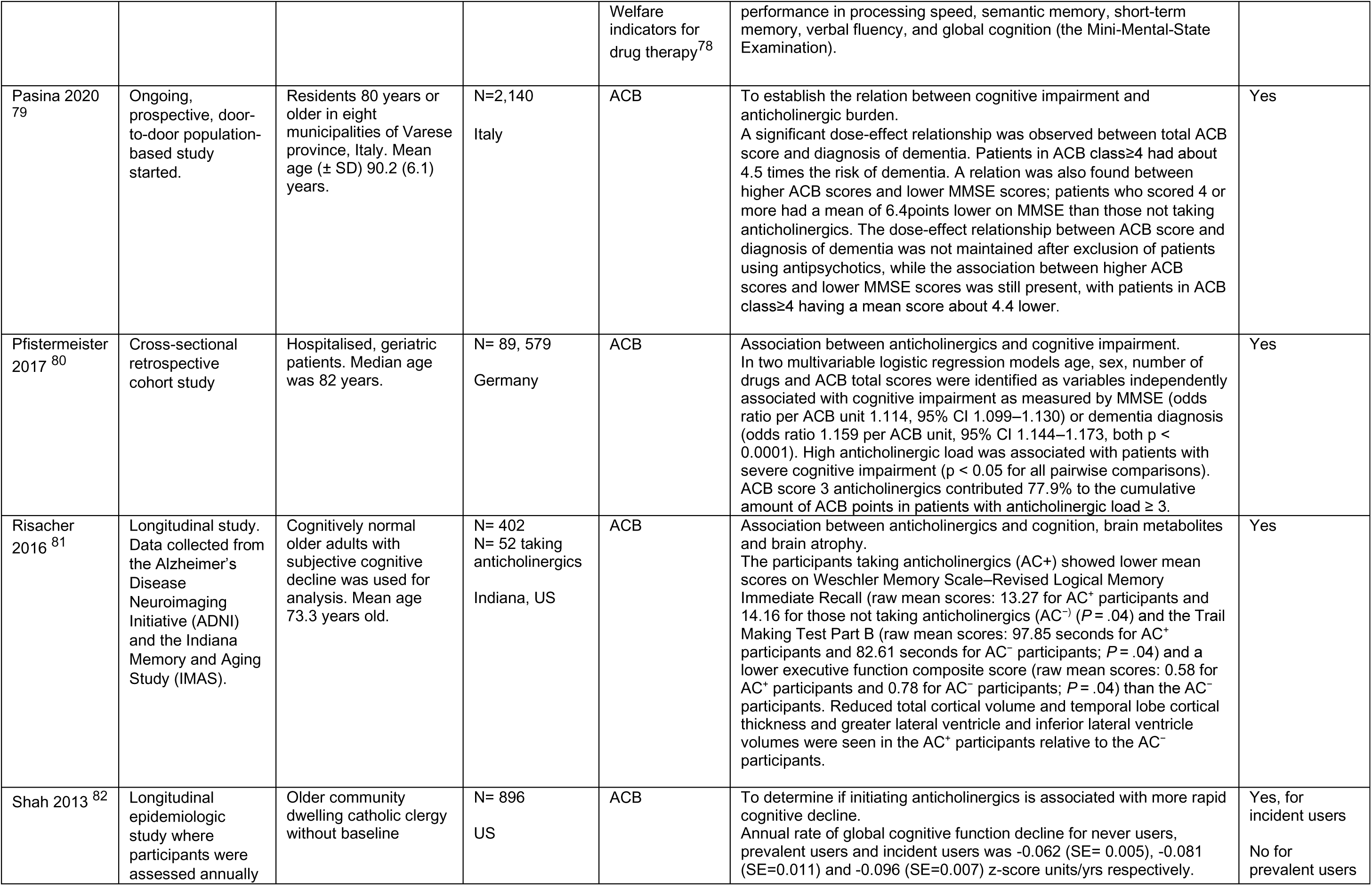
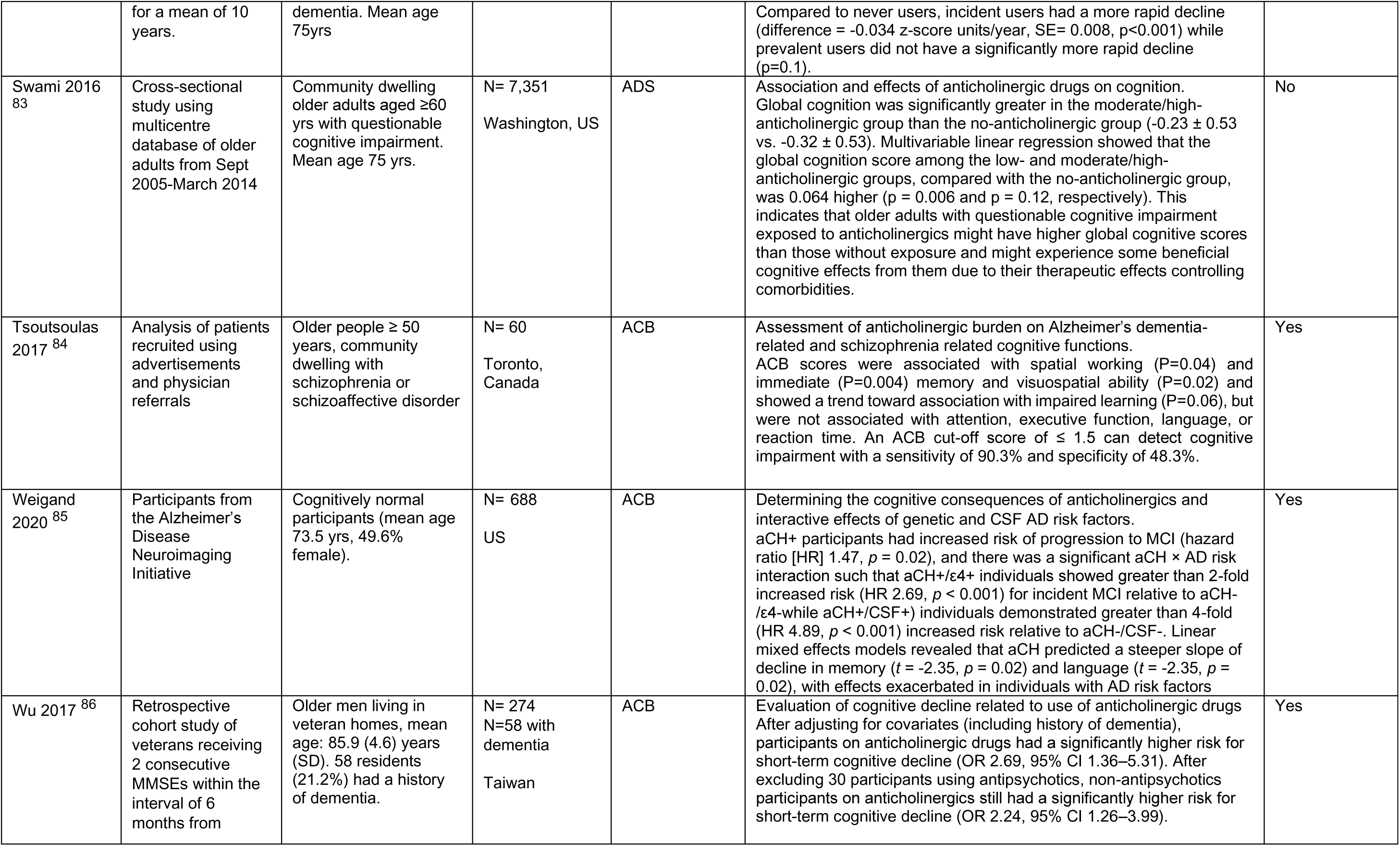
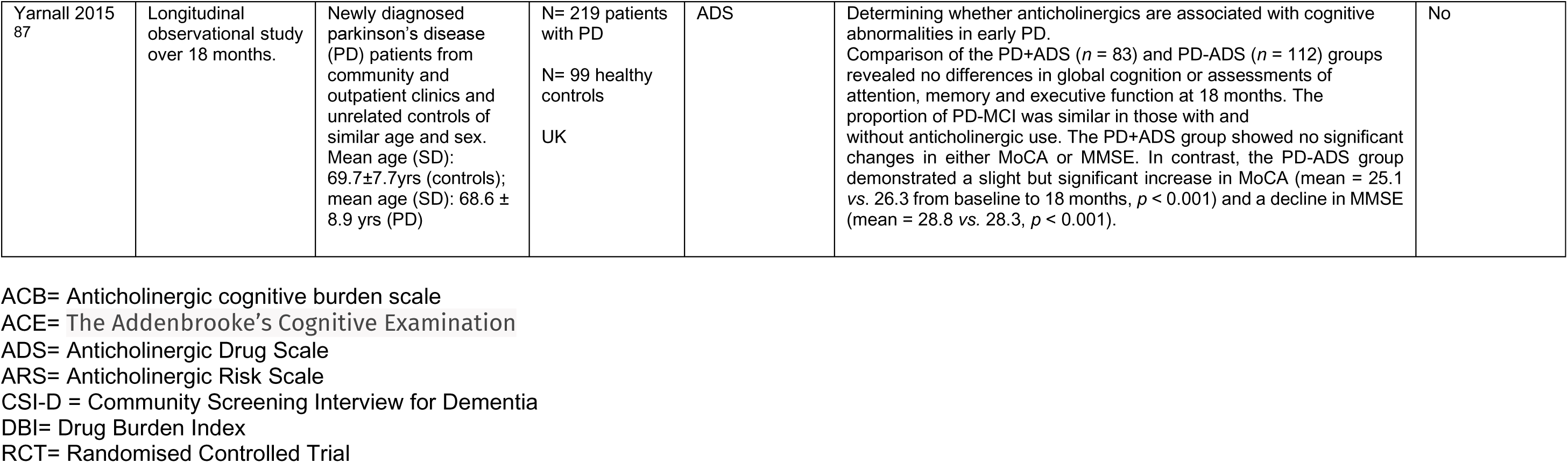
Papers examining the association between anticholinergic drugs and risk of cognitive outcomes.

**Supplementary Table 3.** Papers examining the association between bladder anticholinergic drugs and risk of dementia or cognitive decline.

| Authors/<br>reference | Study Design | Subjects | Sample<br>size/<br>Nation | Anti-<br>cholinergic<br>scale used | Aim/ Main outcomes | Significant<br>association |
| --- | --- | --- | --- | --- | --- | --- |
| Barthold<br>2020 <sup>27</sup> | Retrospective<br>cohort study | Random 20% sample<br>of Medicare<br>beneficiaries using<br>bladder antimuscarinics<br>and ≥ 67 years old. | N= 71,688<br>US | Non-selective:<br>fesoterodine,<br>flavoxate,<br>oxybutynin,<br>tolterodine,<br>tospium.<br>Selective:<br>darifenacin,<br>solifenacin<br>TSDDs | Comparison of Alzheimer's disease and related dementia risk for users of non-selective and M3- selective bladder antimuscarinics. Non-selective antimuscarinic use was not significantly associated with dementia incidence compared to M3 selective drugs. Odds ratios for non-selective use were 0.97 (CI: 0.89-1.04) for 1-364 TSDD, 0.94 (CI: 0.83-1.06) for 365-729, 1.00 (CI: 0.87-1.16) for 730-1094, and 1.03 (CI: 0.88-1.20) for >1094. Higher TSDD of BAMs overall (combining both non-selective and M3-selective BAMs), when compared to 1-364 TSDD, were associated with increased ADRD incidence (OR = 1.05 (CI: 0.99-1.10) for 365-729, OR = 1.11 (CI: 1.05-1.17) for 730-1094, and OR = 1.10 (CI: 1.04-1.15) for >1094). | No difference<br>in dementia<br>risk between<br>selective and<br>non-selective<br>anticholinergics |
| Coupland<br>2019 <sup>2</sup> | Nested case control<br>study in general<br>practice | Patients ≥ 55 yrs<br>registered in GP<br>database. Mean (SD)<br>age of the entire<br>population was 82.2<br>(6.8) yrs. | N= 58,769 with<br>dementia<br>N= 225,574<br>matched<br>controls<br>UK | TSDDs | Evaluation of whether exposure to anticholinergic drugs is associated with dementia risk. Adjusted OR for dementia increased from 1.06 (95% CI, 1.03-1.09) in the lowest anticholinergic exposure category (total exposure of 1-90 TSDDs) to 1.49 (95% CI, 1.44-1.54) in the highest category (>1095 TSDDs), compared with no anticholinergic drug in the 1 to 11 years before the index date. There were significant increases in dementia risk for the bladder antimuscarinic drugs (AOR, 1.65; 95% CI, 1.56-1.75) for more than 1095 TSDDs. | Yes- increased<br>risk of<br>dementia |
| Esin 2015<br>88 | Non-interventional<br>prospective<br>observational study | OAB outpatients. Mean<br>age 73.5 ± 6.1 yrs. Of<br>the 168 patients, 92.3%<br>were female. | N=168<br>Turkey | No<br>Anticholinergic<br>scale used.<br>oxybutynin,<br>darifenacin,<br>tolterodine,<br>trospium, and<br>control groups. | To evaluate effect of antimuscarinic treatment on cognitive functions, depression, and quality of life (QOL) of patients with OAB. MMSE scores were compared, and all groups analysed together, although there was a slight reduction after treatment, this was not statistically significant (p= 0.91). However, in the darifenacin group, there was a slight, but statistically significant decrease in the post-treatment MMSE scores (p= 0.038). Subgroup analyses were performed for patients with dementia and for those with depression. In patients with dementia, no statistically significant MMSE decline was observed (p= 0.72). In patients who have depression, a statistically significant decline in GDS scores was determined (p= 0.01). | Yes – reduced<br>MMSE only for<br>darifenacin |
| Geller 2012<br>89 | Prospective cohort<br>study conducted<br>from Jan to Dec<br>2010, with 12-week<br>follow-up. | Women aged ≥ 55<br>years seeking<br>treatment for OAB.<br>Mean age 70.4 years. | N=35<br>North Carolina,<br>US | Trospium<br>No scale used | To investigate the effect of an anticholinergic medication on cognitive function in postmenopausal women being treated for OAB. Baseline cognitive function was assessed via the Hopkins Verbal Learning Test – Revised Form (HVLT-R) (and its 5 subscales), the Orientation, Memory & Concentration (OMC) short form, and the Mini-Cog evaluation. Cognitive function showed an initial decline on Day 1 in HVLT-R total score (p=0.037), HVLT-R Delayed Recognition subscale (p=0.011) and HVLT-R Recognition Bias subscale (p=0.01). At Week 1 the HVLT-R Learning subscale declined from baseline (p=0.029) but all normalized by Week 4 and remained stable. | No change in<br>cognition<br>(HVLT) for<br>trospium |
| Geller 2017<sup>25</sup> | Randomized double-blind placebo-controlled trial conducted from April 2013 to April 2015. | Women aged ≥50 yrs seeking treatment for OAB randomized to trospium XR 60 mg daily or placebo. Mean age 68yrs. | N=59<br>North Carolina, US | Trospium<br>No scale used | To investigate effect of trospium on cognitive function in postmenopausal women treated for overactive bladder (OAB). There was no difference in HVLT-R total score between trospium and placebo groups at week 4 (P = 0.29). There were also no differences based on the other cognitive tests. | No change in cognition (HVLT) for trospium |
| Gray 2015<sup>47</sup> | Prospective population-based cohort study using data from the Adult Changes in Thought study. | Participants aged ≥ 65 yrs with no dementia, followed up every 2 years. Median age: 74.4 yrs | N= 3,434<br>Seattle, Washington | TSDDs | Examining whether cumulative anticholinergic use is associated with a higher risk for incident dementia. For dementia, adjusted hazard ratios for cumulative anticholinergic use compared with non-use were 0.92 (95% CI, 0.74-1.16) for TDDs of 1 to 90; 1.19 (95% CI, 0.94-1.51) for TSDDs of 91 to 365; 1.23 (95% CI 0.94-1.62) for TSDDs of 366 to 1095, and 1.54 (95% CI, 1.21-1.96) for TSDDs greater than 1095. Higher cumulative anticholinergic medication use is associated with an increased risk for dementia. | Yes- increased risk of dementia |
| Hampel 2017<sup>90</sup> | Observational study | Patients aged ≥70 years prescribed flexible dose of solifenacin for OAB. Mean age 78 ± 6 yrs. | N= 774<br>Germany | Solifenacin<br>No scale used | To evaluate efficacy and safety of solifenacin in older patients with OAB. No change in mean MMSE scores was apparent at week 12. | No change in cognition (MMSE) for solifenacin |
| Iyer 2020<sup>91</sup> | Prospective cohort study | Female patients > 60 years, referred to urogynaecology practice. Mean age 77 years. | N= 106 (N=60 OAB medication, 46 controls)<br>Chicago, US | N=59 oxybutynin<br>N=1 trospium<br>No scale used | To assess cognitive changes in women 12 months after starting anticholinergic medication for OAB. No difference in change of Montreal Cognitive Assessment (MOCA) screening between OAB medication and control groups (0.44 vs 0.56, p=0.646) after controlling for age, depression and ACB score after 12 months of follow-up. | No change in cognition (MOCA) |
| Malcher 2022<sup>28</sup> | Nested case control study using the French National Medical-Administrative Database | Individuals aged >60 years. Controls were matched 5:1 to cases. Median age 82 years. | N=4,810 cases and 24,050 matched controls<br>France | Exposure to flavoxate, oxybutynin, solifenacin, trospium, and fesoterodine defined using daily doses (DDD) | To analyse the relationship between use of anticholinergic drugs to treat OAB and risk of incident dementia. OAB anticholinergic use was associated with an increased risk of dementia (adjusted OR [aOR] [1.23, 95% CI 1.10-1.37] with a cumulative dose response: aOR [1.07 (95% CI 0.91-1.25) for 1-90 DDDs, aOR [1.29 (1.05-1.58) for 91-365 DDDs and aOR [1.48 (1.22-1.80) for >365 DDDs. Considering each drug separately showed a particularly marked increased risk of dementia for oxybutynin and solifenacin, but no increased risk for trospium. | Yes- increased risk of dementia for oxybutynin and solifenacin but not trospium. |
| Matta 2021<sup>29</sup> | Population-based nested case-control study | Patients aged ≥ 66 years with new diagnosis of dementia age and sex matched to controls without dementia | N= 11,392<br>Canada | Antimuscarinic OAB drug or mirabegron<br>No scale used | To differentiate risk of dementia associated with various OAB drugs. Patients receiving solifenacin (OR 1.24; 95% confidence interval 1.08-1.43) and darifenacin (OR 1.30; 95% CI 1.08-1.56) in the prior 6 months had increased odds of incident dementia compared with those receiving mirabegron. In the 6 months to 1 yr prior to diagnosis, receipt of solifenacin (OR 1.34; 95% CI 1.11-1.60), darifenacin (OR 1.49; 95% CI 1.19-1.86), tolterodine (OR 1.21; 95% CI 1.02-1.45), and fesoterodine (OR 1.39; 95% CI 1.14-1.71) was associated with increased odds of | Yes- increased risk of dementia for solifenacin, darifenacin, tolterodine and fesoterodine. |
|  |  |  |  |  | incident dementia compared with receipt of mirabegron. No effect was seen with oxybutynin or trospium. | No for oxybutynin and trospium |
| Moga 2017<br>26 | Retrospective analysis of data from the National Alzheimer's Coordinating Center (NACC) | Cohort age ≥ 65 years and includes not only patients with AD and related disorders but also cognitively normal subjects and those with mild cognitive impairment (MCI). | N= 698 new users<br>N= 7,037 nonusers<br>US | No scale used | Evaluating the association between antimuscarinic initiation and cognitive decline. The odds ratio (OR) and 95% confidence interval for cognitive decline in users as compared to nonusers was 1.4 (1.19–1.65) for Mini- Mental State Examination (MMSE), and 1.21 (1.03–1.42) for Clinical Dementia Rating; in addition, the odds of decline were 20% higher in users compared to nonusers for semantic memory/language and executive function. The effect estimate for MMSE was 1.94 (1.3–2.91) for those with mild cognitive impairment, 1.26 (0.99–1.62) in those with normal cognition, and 1.44 (1.04–1.99) in those with dementia at baseline. | Yes- change in MMSE and Clinical Dementia Rating |
| Richardson 2018<sup>59</sup> | Nested case-control study using GP database (Clinical Practice Research Datalink) | Patients aged 65-99 years with and without dementia. The median age of patients at index date was 83 (interquartile range 78-87) 63% were female. | N= 40, 770 with dementia<br>N= 283, 933 controls without dementia<br>UK | ACB | Association between the duration and level of exposure to different classes of anticholinergic drugs and subsequent incident dementia. When considered by drug class the risk of dementia increased with greater exposure for urological drugs with an ACB score of 3 remained consistently significantly associated with dementia incidence with OR 1.27 95% CI 1.09 to 1.48. Result was also observed for exposure 15-20 years before a diagnosis. | Yes- increased risk of dementia |
| Wagg 2013<br>92 | Twelve-week, randomized, double-blind, placebo-controlled trial | Individuals aged ≥65 yrs (47% male) with OAB symptoms for 3 months or longer. | N= 794<br>Europe, Israel and Turkey | Fesoterodine<br>No scale used | To assess efficacy and safety of fesoterodine in patients with OAB. No meaningful mean change from baseline to week 12 in MMSE score was observed in the fesoterodine (0.24±1.76) or placebo (0.23±1.82) group. Mean MMSE scores at week 12 were 28.4 (range 20–30) in the fesoterodine group and 28.3 (range 19–30) in the placebo group. | No change in MMSE for fesoterodine |
| Wang 2019<br>62 | Taiwan's National Health Insurance Research Database (NHIRD) used to conduct a cohort study | Patients aged ≥ 50 yrs with newly diagnosed lower urinary tract symptoms (LUTS). Mean age was 66.5 yrs, majority were male. | N=16,412<br>Taiwan | Cumulative defined daily doses (cDDD) and ACB | To investigate whether cumulative use of anticholinergics are associated with a higher risk of incident dementia. In a Cox proportional hazards analysis, the adjusted hazard ratio of dementia was 1.15 (95% CI = 0.97–1.37) in the 85– 336 cDDD group, and 1.40 (95% CI = 1.12–1.75) in the ≥337 cDDD group after adjusting for covariates. | Yes- increased risk of dementia |
| Welk 2020<br>93 | Population-based, retrospective, matched cohort study using linked administrative data from Ontario. | Patients with OAB, median age 73 yrs. | N=47,324 users anticholinergics matched to N=23, 662 users of mirabegron<br>Ontario, Canada | No scale used | To determine if there is an increased risk of dementia among patients starting an anticholinergic compared to those starting mirabegron. There was an increased risk of dementia among anticholinergic users compared to mirabegron users (HR: 1.23, 95% CI 1.12–1.35). There was a significant effect modification based on both gender and age; men and those aged ≤75 years on anticholinergics had the highest risk of dementia relative to similar beta-3 agonist users. | Yes- increased risk of dementia |

ACB= Anticholinergic cognitive burden scale
TSDDs= Total standardised daily doses

**Supplementary Table 4.** Papers examining the association between antidepressants with high anticholinergic burden and risk of dementia or cognitive decline.

| Authors/<br>reference | Study Design | Subjects | Sample<br>size/ Nation | Anti-<br>cholinergic<br>scale used | Aim/ Main outcomes | Significant<br>association |
| --- | --- | --- | --- | --- | --- | --- |
| Bishara<br>2021 <sup>39</sup> | Retrospective cohort<br>study | Patients with dementia ≥<br>50 years taking<br>antidepressants. | N=4380<br>UK | AEC | To investigate associations between antidepressants or<br>antipsychotics and mortality, hospitalisation and cognitive decline in<br>people with dementia.<br>There was reduced mortality risk in people receiving antidepressants<br>with high central anticholinergic burden compared to those with no or<br>low burden (Hazard ratio (HR): 0.88; 95% confidence interval (CI):<br>0.79-0.98; p=0.023). No significant associations were detected<br>between antidepressant's central anticholinergic burden and<br>hospitalisation or cognitive decline. | No association<br>for cognitive<br>decline |
| Coupland<br>2019 <sup>2</sup> | Nested case control<br>study in general<br>practice | Patients ≥ 55 yrs<br>registered in GP<br>database. Mean (SD)<br>age of the entire<br>population was 82.2<br>(6.8) yrs. | N= 58,769 with<br>dementia<br>N= 225,574<br>matched<br>controls<br>UK | TSDDs | Evaluation of whether exposure to anticholinergic drugs is associated<br>with dementia risk. Adjusted OR for dementia increased from 1.06<br>(95% CI, 1.03-1.09) in the lowest anticholinergic exposure category<br>(total exposure of 1-90 TSDDs) to 1.49 (95% CI, 1.44-1.54) in the<br>highest category (>1095 TSDDs), compared with no anticholinergic<br>drug in the 1 to 11 years before the index date. There were<br>significant increases in dementia risk for anticholinergic<br>antidepressants (adjusted OR [AOR], 1.29; 95% CI, 1.24-1.34) for<br>more than 1095 TSDDs. | Yes- increased<br>risk of dementia |
| Heath 2018<br><sup>37</sup> | Prospective cohort<br>study using an<br>integrated<br>healthcare delivery<br>system. | Community-dwelling<br>individuals aged ≥65 yrs<br>without dementia. Mean<br>age 75.2±6.3. Mean<br>follow up 7.7 yrs. | N= 3,059<br>Washington,<br>US | TDDs | Antidepressant use and associated dementia risk.<br>Individual antidepressant classes were not associated with<br>differences in dementia risk, although paroxetine use was associated<br>with higher risk of dementia for all TSDD categories than no use (0–<br>90 TSDDs: hazard ratio (HR)=1.69, 95% confidence interval<br>(CI)=1.18–2.42; 91–365 TSDDs: HR=1.40, 95% CI=0.88–2.23; 366–<br>1095 TSDDs: HR=2.13, 95% CI=1.32–3.43; ≥1095 TSDDs:<br>HR=1.42, 95% CI=0.82–2.46). | Increased risk<br>of dementia<br>only for<br>paroxetine |
| Heser 2018<br><sup>38</sup> | Prospective multi-<br>centre cohort study<br>with follow-up up to<br>12 years | Participants at least 75<br>years old in primary care<br>were recruited via GPs | N= 3,239<br>Germany | Priscus list by<br>Holt et al.<br>2010 | Investigated whether intake of antidepressants considered potentially<br>inappropriate medication (PIM) according to Priscus list would predict<br>incident dementia.<br>PIM antidepressants (HR = 1.49, 95% CI: 1.06-2.10, p = .021), but<br>not other antidepressants (HR = 1.04, 95% CI: 0.66-1.66, p = .863),<br>were associated with increased risk for subsequent dementia (in age,<br>sex-, education-, and depressive symptoms adjusted models).<br>Significant associations disappeared after global cognition at<br>baseline was controlled for. | Yes increased<br>risk of dementia<br>for PIM<br>antidepressants |
| Mate 2022<br>40 | Cross-sectional study using a structural interview process | Aboriginal communities aged ≥ 60 years. Mean age 66.6 years, 59.5% female. | N=336<br>New South Wales, Australia | ACB | Examination of medication use, anticholinergic burden among aboriginal people. When adjusted for sex, age and number of comorbidities anticholinergic burden was not associated with cognitive impairment, falls or hospitalisation during the past 12 months but was still associated with depressive symptoms (OR 2.86; 95% CI 1.48-5.51). | No association with cognitive impairment, falls or hospitalisation |
| Richardson 2018 <sup>59</sup> | Nested case-control study using GP database (Clinical Practice Research Datalink) | Patients aged 65-99 years with and without dementia. Median age 83 years 63% were female. | N= 40, 770 (dementia)<br>N=283, 933 (controls)<br>UK | ACB | Association between the duration and level of exposure to different classes of anticholinergic drugs and subsequent incident dementia. Antidepressant drugs with an ACB score of 3 remained consistently significantly associated with dementia incidence with OR 1.19 95% CI 1.10 to 1.29. | Yes- increased risk of dementia |
| Saczynski 2015 <sup>41</sup> | Data drawn from the Health and Retirement (HRS) Study. Longitudinal survey- 6 year follow up | Participants aged ≥ 50 years from HRS who had self-reported antidepressant use. | N= 3714<br>US | No anticholinergic scale used | Assessment of the association between antidepressant use and cognitive decline (battery of tests) over 6 years. During the follow up, cognition declined in both users and nonusers of antidepressants, ranging from -1.4 change in mean score in those with high depressive symptoms taking antidepressants to -0.5 change in mean score in those with high depressive symptoms not taking antidepressants. In adjusted models, cognition declined in those on antidepressants at the same rate as those not taking them. Results remained consistent across different levels of baseline cognitive function, age, duration of use (prolonged vs short). | No change in cognitive function |

AEC= Anticholinergic Effect on Cognition scale
ACB= Anticholinergic cognitive burden scale
TSDDs= Total standardised daily doses

TSDDs= Total standardised daily doses
| Section and Topic | Item # | Checklist item | Location where item is reported |
| --- | --- | --- | --- |
| **TITLE** |  |  |  |
| Title | 1 | Identify the report as a systematic review. | Title |
| **ABSTRACT** |  |  |  |
| Abstract | 2 | See the PRISMA 2020 for Abstracts checklist. | Abstract |
| **INTRODUCTION** |  |  |  |
| Rationale | 3 | Describe the rationale for the review in the context of existing knowledge. | Intro |
| Objectives | 4 | Provide an explicit statement of the objective(s) or question(s) the review addresses. | Intro |
| **METHODS** |  |  |  |
| Eligibility criteria | 5 | Specify the inclusion and exclusion criteria for the review and how studies were grouped for the syntheses. | Methods |
| Information sources | 6 | Specify all databases, registers, websites, organisations, reference lists and other sources searched or consulted to identify studies. Specify the date when each source was last searched or consulted. | Fig 1 |
| Search strategy | 7 | Present the full search strategies for all databases, registers and websites, including any filters and limits used. | Fig 1 |
| Selection process | 8 | Specify the methods used to decide whether a study met the inclusion criteria of the review, including how many reviewers screened each record and each report retrieved, whether they worked independently, and if applicable, details of automation tools used in the process. | Methods |
| Data collection process | 9 | Specify the methods used to collect data from reports, including how many reviewers collected data from each report, whether they worked independently, any processes for obtaining or confirming data from study investigators, and if applicable, details of automation tools used in the process. | Methods |
| Data items | 10a | List and define all outcomes for which data were sought. Specify whether all results that were compatible with each outcome domain in each study were sought (e.g. for all measures, time points, analyses), and if not, the methods used to decide which results to collect. | Methods |
|  | 10b | List and define all other variables for which data were sought (e.g. participant and intervention characteristics, funding sources). Describe any assumptions made about any missing or unclear information. | Methods |
| Study risk of bias assessment | 11 | Specify the methods used to assess risk of bias in the included studies, including details of the tool(s) used, how many reviewers assessed each study and whether they worked independently, and if applicable, details of automation tools used in the process. | Methods |
| Effect measures | 12 | Specify for each outcome the effect measure(s) (e.g. risk ratio, mean difference) used in the synthesis or presentation of results. | Not limited |
| Synthesis methods | 13a | Describe the processes used to decide which studies were eligible for each synthesis (e.g. tabulating the study intervention characteristics and comparing against the planned groups for each synthesis (item #5)). | Results – no synthesis possible |
|  | 13b | Describe any methods required to prepare the data for presentation or synthesis, such as handling of missing summary statistics, or data conversions. | NA |
|  | 13c | Describe any methods used to tabulate or visually display results of individual studies and syntheses. | NA |
|  | 13d | Describe any methods used to synthesize results and provide a rationale for the choice(s). If meta-analysis was performed, describe the model(s), method(s) to identify the presence and extent of statistical heterogeneity, and software package(s) used. | NA |
|  | 13e | Describe any methods used to explore possible causes of heterogeneity among study results (e.g. subgroup analysis, meta-regression). | NA |
|  | 13f | Describe any sensitivity analyses conducted to assess robustness of the synthesized results. | NA |
| Reporting bias assessment | 14 | Describe any methods used to assess risk of bias due to missing results in a synthesis (arising from reporting biases). | NA |
| Certainty assessment | 15 | Describe any methods used to assess certainty (or confidence) in the body of evidence for an outcome. | No formal – see discussion |
| **RESULTS** |  |  |  |
| Study selection | 16a | Describe the results of the search and selection process, from the number of records identified in the search to the number of studies included in the review, ideally using a flow diagram. | Fig 1 |
|  | 16b | Cite studies that might appear to meet the inclusion criteria, but which were excluded, and explain why they were excluded. | supplementary |
| Study characteristics | 17 | Cite each included study and present its characteristics. | tables |
| Risk of bias in studies | 18 | Present assessments of risk of bias for each included study. | Result text |
| Results of individual studies | 19 | For all outcomes, present, for each study: (a) summary statistics for each group (where appropriate) and (b) an effect estimate and its precision (e.g. confidence/credible interval), ideally using structured tables or plots. | Fig 2 |
| Results of syntheses | 20a | For each synthesis, briefly summarise the characteristics and risk of bias among contributing studies. | NA |
|  | 20b | Present results of all statistical syntheses conducted. If meta-analysis was done, present for each the summary estimate and its precision (e.g. confidence/credible interval) and measures of statistical heterogeneity. If comparing groups, describe the direction of the effect. | NA |
|  | 20c | Present results of all investigations of possible causes of heterogeneity among study results. | NA |
|  | 20d | Present results of all sensitivity analyses conducted to assess the robustness of the synthesized results. | NA |
| Reporting biases | 21 | Present assessments of risk of bias due to missing results (arising from reporting biases) for each synthesis assessed. | NA |
| Certainty of evidence | 22 | Present assessments of certainty (or confidence) in the body of evidence for each outcome assessed. | No formal – see discussion |
| **DISCUSSION** |  |  |  |
| Discussion | 23a | Provide a general interpretation of the results in the context of other evidence. | Discussion |
|  | 23b | Discuss any limitations of the evidence included in the review. | Discussion |
|  | 23c | Discuss any limitations of the review processes used. | Discussion |
|  | 23d | Discuss implications of the results for practice, policy, and future research. | Discussion |
| **OTHER INFORMATION** |  |  |  |
| Registration and protocol | 24a | Provide registration information for the review, including register name and registration number, or state that the review was not registered. | Not registered |
|  | 24b | Indicate where the review protocol can be accessed, or state that a protocol was not prepared. | Not registered |
|  | 24c | Describe and explain any amendments to information provided at registration or in the protocol. | NA |
| Support | 25 | Describe sources of financial or non-financial support for the review, and the role of the funders or sponsors in the review. | Funding |
| Competing interests | 26 | Declare any competing interests of review authors. | Disclosure statement |
| Availability of data, code and other materials | 27 | Report which of the following are publicly available and where they can be found: template data collection forms; data extracted from included studies; data used for all analyses; analytic code; any other materials used in the review. | None produced |

From: Page MJ, McKenzie JE, Bossuyt PM, Boutron I, Hoffmann TC, Mulrow CD, et al. The PRISMA 2020 statement: an updated guideline for reporting systematic reviews. BMJ 2021;372:n71. doi: 10.1136/bmj.n71

## References

1. Campbell NL, Boustani MA, Lane KA, et al. Use of anticholinergics and the risk of cognitive impairment in an African American population. Neurology 2010;75(2):152–59. doi: 10.1212/WNL.0b013e3181e7f2ab

2. Coupland CAC, Hill T, Dening T, et al. Anticholinergic Drug Exposure and the Risk of Dementia: A Nested Case-Control Study. JAMA Internal Medicine 2019;179(8):1084–93. doi: 10.1001/jamainternmed.2019.0677 %J JAMA Internal Medicine

3. Richardson K, Fox C, Maidment I, et al. Anticholinergic drugs and risk of dementia: case-control study. 2018;361:k1315. doi: 10.1136/bmj.k1315 %J BMJ

4. McMichael AJ, Zafeiridi E, Ryan M, et al. Anticholinergic drug use and risk of mortality for people with dementia in Northern Ireland. Aging & Mental Health 2021;25(8):1475–82. doi: 10.1080/13607863.2020.1830028

5. Dmochowski RR, Thai S, Iglay K, et al. Increased risk of incident dementia following use of anticholinergic agents: A systematic literature review and meta-analysis. Neurourol Urodyn 2021;40(1):28–37. doi: 10.1002/nau.24536 [published Online First: 2020/10/25]

6. Zheng YB, Shi L, Zhu XM, et al. Anticholinergic drugs and the risk of dementia: A systematic review and meta-analysis. Neuroscience and biobehavioral reviews 2021;127:296–306. doi: 10.1016/j.neubiorev.2021.04.031 [published Online First: 2021/05/03]

7. Fox C, Smith T, Maidment I, et al. Effect of medications with anti-cholinergic properties on cognitive function, delirium, physical function and mortality: a systematic review. Age Ageing 2014;43(5):604–15. doi: 10.1093/ageing/afu096 [published Online First: 20140719]

8. Andre L, Gallini A, Montastruc F, et al. Association between anticholinergic (atropinic) drug exposure and cognitive function in longitudinal studies among individuals over 50 years old: a systematic review. Eur J Clin Pharmacol 2019;75(12):1631–44. doi: 10.1007/s00228-019-02744-8 [published Online First: 2019/08/31]

9. Pieper NT, Grossi CM, Chan WY, et al. Anticholinergic drugs and incident dementia, mild cognitive impairment and cognitive decline: a meta-analysis. Age Ageing 2020;49(6):939–47. doi: 10.1093/ageing/afaa090 [published Online First: 2020/07/01]

10. Ah Y-M, Suh Y, Jun K, et al. Effect of anticholinergic burden on treatment modification, delirium and mortality in newly diagnosed dementia patients starting a cholinesterase inhibitor: A population-based study. Basic & Clinical Pharmacology & Toxicology 2019;124(6):741–48. doi: 10.1111/bcpt.13184

11. Bishara D, Perera G, Harwood D, et al. The anticholinergic effect on cognition (AEC) scale— Associations with mortality, hospitalisation and cognitive decline following dementia diagnosis. International Journal of Geriatric Psychiatry 2020;35(9):1069–77. doi: 10.1002/gps.5330

12. Gnjidic D, Hilmer SN, Hartikainen S, et al. Impact of High Risk Drug Use on Hospitalization and Mortality in Older People with and without Alzheimer’s Disease: A National Population Cohort Study. PloS one 2014;9(1):e83224. doi: 10.1371/journal.pone.0083224

13. Tan E, Eriksdotter M, García-Ptacek S, et al. Anticholinergic Burden and Risk of Stroke and Death in People with Different Types of Dementia. Journal of Alzheimer’s Disease 2018;65:1–8. doi: 10.3233/JAD-180353

14. Cross AJ, George J, Woodward MC, et al. Potentially Inappropriate Medication, Anticholinergic Burden, and Mortality in People Attending Memory Clinics. Journal of Alzheimer’s disease : JAD 2017;60(2):349–58. doi: 10.3233/jad-170265 [published Online First: 2017/09/05]

15. Buckley E, Jonsson A, Flood Z, et al. Potentially inappropriate medication use and mortality in patients with cognitive impairment. Eur J Clin Pharmacol 2022;78(12):2013–20. doi: 10.1007/s00228-022-03410-2 [published Online First: 20221103]

16. Tan ECK, Eriksdotter M, Garcia-Ptacek S, et al. Anticholinergic Burden and Risk of Stroke and Death in People with Different Types of Dementia. Journal of Alzheimer’s disease : JAD 2018;65(2):589–96. doi: 10.3233/jad-180353 [published Online First: 2018/07/30]

17. Tan ECK, Eriksdotter M, Garcia-Ptacek S, et al. Anticholinergic Burden and Risk of Stroke and Death in People with Different Types of Dementia. Journal of Alzheimer’s Disease 2018;65:589–96. doi: 10.3233/JAD-180353

18. Wang K, Alan J, Page AT, et al. Anticholinergics and clinical outcomes amongst people with pre-existing dementia: A systematic review. Maturitas 2021;151:1–14. doi: 10.1016/j.maturitas.2021.06.004 [published Online First: 20210620]

19. Watanabe S, Fukatsu T, Kanemoto K. Risk of hospitalization associated with anticholinergic medication for patients with dementia. Psychogeriatrics 2018;18(1):57–63. doi: 10.1111/psyg.12291

20. Liang C-K, Chou M-Y, Hsu Y-H, et al. The association of potentially inappropriate medications, polypharmacy and anticholinergic burden with readmission and emergency room revisit after discharge: A hospital-based retrospective cohort study. British Journal of Clinical Pharmacology 2023;89(1):187–200. doi: 10.1111/bcp.15457

21. Dyer AH, Murphy C, Segurado R, et al. Is Ongoing Anticholinergic Burden Associated With Greater Cognitive Decline and Dementia Severity in Mild to Moderate Alzheimer’s Disease? The journals of gerontology Series A, Biological sciences and medical sciences 2020;75(5):987–94. doi: 10.1093/gerona/glz244 [published Online First: 2019/10/16]

22. Fox C, Livingston G, Maidment ID, et al. The impact of anticholinergic burden in Alzheimer’s Dementia-the Laser-AD study. Age and Ageing 2011;40(6):730–35. doi: 10.1093/ageing/afr102 %J Age and Ageing

23. Lampela P, Lavikainen P, Garcia-Horsman JA, et al. Anticholinergic Drug Use, Serum Anticholinergic Activity, and Adverse Drug Events Among Older People: A Population-Based Study. Drugs & aging 2013;30(5):321–30. doi: 10.1007/s40266-013-0063-2

24. Sanders LMJ, Hortobágyi T, van Staveren G, et al. Relationship between drug burden and physical and cognitive functions in a sample of nursing home patients with dementia. Eur J Clin Pharmacol 2017;73(12):1633–42. doi: 10.1007/s00228-017-2319-y [published Online First: 2017/09/19]

25. Geller EJ, Dumond JB, Bowling JM, et al. Effect of Trospium Chloride on Cognitive Function in Women Aged 50 and Older: A Randomized Trial. Female pelvic medicine & reconstructive surgery 2017;23(2):118–23. doi: 10.1097/spv.0000000000000374 [published Online First: 2017/01/10]

26. Moga DC, Abner EL, Wu Q, et al. Bladder antimuscarinics and cognitive decline in elderly patients. Alzheimers Dement (N Y) 2017;3(1):139–48. doi: 10.1016/j.trci.2017.01.003

27. Barthold D, Marcum ZA, Gray SL, et al. Alzheimer’s disease and related dementias risk: Comparing users of non-selective and M3-selective bladder antimuscarinic drugs. Pharmacoepidemiology and Drug Safety 2020;29(12):1650–58. doi: 10.1002/pds.5098

28. Malcher MF, Droupy S, Berr C, et al. Dementia Associated with Anticholinergic Drugs Used for Overactive Bladder: A Nested Case-Control Study Using the French National Medical-Administrative Database. Journal of Urology 2022;208(4):863–71. doi: doi:10.1097/JU.0000000000002804

29. Matta R, Gomes T, Juurlink D, et al. Receipt of Overactive Bladder Drugs and Incident Dementia: A Population-based Case-control Study. Eur Urol Focus 2021 doi: 10.1016/j.euf.2021.10.009 [published Online First: 2021/11/08]

30. Triantafylidis LK, Clemons JS, Peron EP, et al. Brain Over Bladder: A Systematic Review of Dual Cholinesterase Inhibitor and Urinary Anticholinergic Use. Drugs & aging 2018;35(1):27–41. doi: 10.1007/s40266-017-0510-6

31. Paquette A, Gou P, Tannenbaum C. Systematic Review and Meta-Analysis: Do Clinical Trials Testing Antimuscarinic Agents for Overactive Bladder Adequately Measure Central Nervous System Adverse Events? J Am Geriatr Soc 2011;59(7):1332–39. doi: 10.1111/j.1532-5415.2011.03473.x

32. Rangganata E, Widia F, Rahardjo HE. Effect of Antimuscarinic Drugs on Cognitive Functions in the Management of Overactive Bladder in Elderly. Acta medica Indonesiana 2020;52 3:255–63.

33. McMichael AJ, Zafeiridi E, Ryan M, et al. Anticholinergic drug use and risk of mortality for people with dementia in Northern Ireland. Aging & Mental Health 2020:1–8. doi: 10.1080/13607863.2020.1830028

34. Kachru N, Holmes HM, Johnson ML, et al. Risk of Mortality Associated with Non-selective Antimuscarinic medications in Older Adults with Dementia: a Retrospective Study. Journal of general internal medicine 2020;35(7):2084–93. doi: 10.1007/s11606-020-05634-3 [published Online First: 2020/02/05]

35. Bishara D, Perera G, Harwood D, et al. Centrally Acting Anticholinergic Drugs Used for Urinary Conditions Associated with Worse Outcomes in Dementia. Journal of the American Medical Directors Association 2021;22(12):2547–52. doi: 10.1016/j.jamda.2021.08.011 [published Online First: 20210830]

36. Kachru N, Holmes HM, Johnson ML, et al. Comparative risk of adverse outcomes associated with nonselective and selective antimuscarinic medications in older adults with dementia and overactive bladder. International journal of geriatric psychiatry 2021;36(5):684–96. doi: 10.1002/gps.5467

37. Heath L, Gray SL, Boudreau DM, et al. Cumulative Antidepressant Use and Risk of Dementia in a Prospective Cohort Study. J Am Geriatr Soc 2018;66(10):1948–55. doi: 10.1111/jgs.15508 [published Online First: 2018/09/18]

38. Heser K, Luck T, Röhr S, et al. Potentially inappropriate medication: Association between the use of antidepressant drugs and the subsequent risk for dementia. J Affect Disord 2018;226:28–35. doi: 10.1016/j.jad.2017.09.016 [published Online First: 2017/09/25]

39. Bishara D, Perera G, Harwood D, et al. Centrally-acting anticholinergic drugs– associations with mortality, hospitalisation and cognitive decline following dementia diagnosis in people receiving antidepressant and antipsychotic drugs. Aging & Mental Health 2021:1–9. doi: 10.1080/13607863.2021.1947967

40. Mate K, Kerr K, Priestley A, et al. Use of tricyclic antidepressants and other anticholinergic medicines by older Aboriginal Australians: association with negative health outcomes. International Psychogeriatrics 2022;34(1):71–78. doi: 10.1017/S104161022000174X [published Online First: 2020/09/28]

41. Saczynski JS, Rosen AB, McCammon RJ, et al. Antidepressant Use and Cognitive Decline: The Health and Retirement Study. The American journal of medicine 2015;128(7):739–46. doi: 10.1016/j.amjmed.2015.01.007 [published Online First: 20150130]

42. Monroe T, Carter M. Using the Folstein Mini Mental State Exam (MMSE) to explore methodological issues in cognitive aging research. Eur J Ageing 2012;9(3):265–74. doi: 10.1007/s10433-012-0234-8 [published Online First: 20120615]

43. Doraiswamy PM, Krishnan KRR, Oxman T, et al. Does Antidepressant Therapy Improve Cognition in Elderly Depressed Patients? The Journals of Gerontology: Series A 2003;58(12):M1137–M44. doi: 10.1093/gerona/58.12.M1137

44. Cai X, Campbell N, Khan B, et al. Long-term anticholinergic use and the aging brain. Alzheimer’s & dementia : the journal of the Alzheimer’s Association 2013;9(4):377–85. doi: 10.1016/j.jalz.2012.02.005 [published Online First: 2012/11/28]

45. Chatterjee S, Bali V, Carnahan RM, et al. Anticholinergic Medication Use and Risk of Dementia Among Elderly Nursing Home Residents with Depression. The American journal of geriatric psychiatry : official journal of the American Association for Geriatric Psychiatry 2016;24(6):485–95. doi: 10.1016/j.jagp.2015.12.011 [published Online First: 2016/03/16]

46. Gildengers A, Stoehr GP, Ran X, et al. Anticholinergic Drug Burden and Risk of Incident MCI and Dementia: A Population-based Study. Alzheimer Disease & Associated Disorders 2023;37(1)

47. Gray SL, Anderson ML, Dublin S, et al. Cumulative use of strong anticholinergics and incident dementia: a prospective cohort study. JAMA Intern Med 2015;175(3):401–7. doi: 10.1001/jamainternmed.2014.7663 [published Online First: 2015/01/27]

48. Grossi CM, Richardson K, Fox C, et al. Anticholinergic and benzodiazepine medication use and risk of incident dementia: a UK cohort study. BMC geriatrics 2019;19(1):276. doi: 10.1186/s12877-019-1280-2 [published Online First: 2019/10/23]

49. Hafdi M, Hoevenaar-Blom MP, Beishuizen CRL, et al. Association of Benzodiazepine and Anticholinergic Drug Usage With Incident Dementia: A Prospective Cohort Study of Community-Dwelling Older Adults. Journal of the American Medical Directors Association 2020;21(2):188–93.e3. doi: 10.1016/j.jamda.2019.05.010 [published Online First: 2019/07/14]

50. Hsu WH, Wen YW, Chen LK, et al. Comparative Associations Between Measures of Anti-cholinergic Burden and Adverse Clinical Outcomes. Ann Fam Med 2017;15(6):561–69. doi: 10.1370/afm.2131 [published Online First: 2017/11/15]

51. Hsu W-H, Huang S-T, Lu W-H, et al. Impact of Multiple Prescriptions With Anticholinergic Properties on Adverse Clinical Outcomes in the Elderly: A Longitudinal Cohort Study in Taiwan. Clinical Pharmacology & Therapeutics 2021;110(4):966–74. doi: 10.1002/cpt.2217

52. Jessen F, Kaduszkiewicz H, Daerr M, et al. Anticholinergic drug use and risk for dementia: target for dementia prevention. Eur Arch Psychiatry Clin Neurosci 2010;260 Suppl 2:S111–5. doi: 10.1007/s00406-010-0156-4 [published Online First: 2010/10/21]

53. Joung KI, Kim S, Cho YH, et al. Association of Anticholinergic Use with Incidence of Alzheimer’s Disease: Population-based Cohort Study. Sci Rep 2019;9(1):6802. doi: 10.1038/s41598-019-43066-0 [published Online First: 2019/05/03]

54. Liu YP, Chien WC, Chung CH, et al. Are Anticholinergic Medications Associated With Increased Risk of Dementia and Behavioral and Psychological Symptoms of Dementia? A Nationwide 15-Year Follow-Up Cohort Study in Taiwan. Front Pharmacol 2020;11:30. doi: 10.3389/fphar.2020.00030 [published Online First: 2020/03/03]

55. Lockery JE, Broder JC, Ryan J, et al. A Cohort Study of Anticholinergic Medication Burden and Incident Dementia and Stroke in Older Adults. Journal of General Internal Medicine 2021;36(6):1629–37. doi: 10.1007/s11606-020-06550-2

56. Mur J, Russ TC, Cox SR, et al. Association between anticholinergic burden and dementia in UK Biobank. Alzheimer’s & Dementia: Translational Research & Clinical Interventions 2022;8(1):e12290. doi: 10.1002/trc2.12290

57. Park H-Y, Park J-W, Song HJ, et al. The Association between Polypharmacy and Dementia: A Nested Case-Control Study Based on a 12-Year Longitudinal Cohort Database in South Korea. PloS one 2017;12(1):e0169463. doi: 10.1371/journal.pone.0169463

58. By the American Geriatrics Society Beers Criteria Update Expert P. American Geriatrics Society 2015 Updated Beers Criteria for Potentially Inappropriate Medication Use in Older Adults. J Am Geriatr Soc 2015;63(11):2227–46. doi: 10.1111/jgs.13702

59. Richardson K, Fox C, Maidment I, et al. Anticholinergic drugs and risk of dementia: case-control study. BMJ 2018;361:k1315. doi: 10.1136/bmj.k1315

60. Suh Y, Ah YM, Han E, et al. Dose response relationship of cumulative anticholinergic exposure with incident dementia: validation study of Korean anticholinergic burden scale. BMC geriatrics 2020;20(1):265. doi: 10.1186/s12877-020-01671-z [published Online First: 2020/07/31]

61. Trevisan C, Limongi F, Siviero P, et al. Mild polypharmacy and MCI progression in older adults: the mediation effect of drug–drug interactions. Aging Clinical and Experimental Research 2021;33(1):49–56. doi: 10.1007/s40520-019-01420-2

62. Wang Y-C, Chen Y-L, Huang C-C, et al. Cumulative use of therapeutic bladder anticholinergics and the risk of dementia in patients with lower urinary tract symptoms: a nationwide 12-year cohort study. BMC geriatrics 2019;19(1):380. doi: 10.1186/s12877-019-1401-y

63. Wilczyński K, Gorczyca M, Gołębiowska J, et al. Anticholinergic Burden of Geriatric Ward Inpatients. Medicina (Kaunas) 2021;57(10):1115. doi: 10.3390/medicina57101115

64. Andre L, Gallini A, Montastruc F, et al. Anticholinergic exposure and cognitive decline in older adults: effect of anticholinergic exposure definitions in a 3-year analysis of the multidomain Alzheimer preventive trial (MAPT) study. Br J Clin Pharmacol 2019;85(1):71–99. doi: 10.1111/bcp.13734 [published Online First: 2018/08/12]

65. Broder JC, Ryan J, Shah RC, et al. Anticholinergic medication burden and cognitive function in participants of the ASPREE study. Pharmacotherapy 2022;42(2):134–44. doi: 10.1002/phar.2652 [published Online First: 2021/12/14]

66. Campbell NL, Perkins AJ, Bradt P, et al. Association of Anticholinergic Burden with Cognitive Impairment and Health Care Utilization Among a Diverse Ambulatory Older Adult Population. Pharmacotherapy: The Journal of Human Pharmacology and Drug Therapy 2016;36(11):1123–31. doi: 10.1002/phar.1843

67. Campbell NL, Lane KA, Gao S, et al. Anticholinergics Influence Transition from Normal Cognition to Mild Cognitive Impairment in Older Adults in Primary Care. Pharmacotherapy 2018;38(5):511–19. doi: 10.1002/phar.2106 [published Online First: 2018/03/31]

68. Chatterjee S, Bali V, Carnahan RM, et al. Anticholinergic burden and risk of cognitive impairment in elderly nursing home residents with depression. Research in Social and Administrative Pharmacy 2020;16(3):329–35. doi: 10.1016/j.sapharm.2019.05.020

69. Dauphinot V, Mouchoux C, Veillard S, et al. Anticholinergic drugs and functional, cognitive impairment and behavioral disturbances in patients from a memory clinic with subjective cognitive decline or neurocognitive disorders. Alzheimers Res Ther 2017;9(1):58–58. doi: 10.1186/s13195-017-0284-4

70. Dyer AH, Laird E, Hoey L, et al. Long-term anticholinergic, benzodiazepine and Z-drug use in community-dwelling older adults: What is the impact on cognitive and neuropsychological performance? International journal of geriatric psychiatry 2021;36(11):1767–77. doi: 10.1002/gps.5598

71. Gnjidic D, Le Couteur DG, Naganathan V, et al. Effects of Drug Burden Index on Cognitive Function in Older Men. Journal of Clinical Psychopharmacology 2012;32(2)

72. Grande G, Tramacere I, Vetrano DL, et al. Role of anticholinergic burden in primary care patients with first cognitive complaints. Eur J Neurol 2017;24(7):950–55. doi: 10.1111/ene.13313 [published Online First: 2017/05/16]

73. Jamsen KM, Gnjidic D, Hilmer SN, et al. Drug Burden Index and change in cognition over time in community-dwelling older men: the CHAMP study. Annals of Medicine 2017;49(2):157–64. doi: 10.1080/07853890.2016.1252053

74. Koyama A, Steinman M, Ensrud K, et al. Long-term cognitive and functional effects of potentially inappropriate medications in older women. The journals of gerontology Series A, Biological sciences and medical sciences 2014;69(4):423–9. doi: 10.1093/gerona/glt192 [published Online First: 2013/12/03]

75. Kusljic S, Woolley A, Lowe M, et al. How do cognitive and functional impairment relate to the use of anticholinergic medications in hospitalised patients aged 65 years and over? Aging Clinical and Experimental Research 2020;32(3):423–31. doi: 10.1007/s40520-019-01225-3

76. López-Matons N, Conill Badell D, Obrero Cusidó G, et al. Anticholinergic drugs and cognitive impairment in the elderly. Med Clin (Barc) 2018;151(4):141–44. doi: 10.1016/j.medcli.2018.01.014 [published Online First: 2018/03/12]

77. Papenberg G, Bäckman L, Fratiglioni L, et al. Anticholinergic drug use is associated with episodic memory decline in older adults without dementia. Neurobiology of Aging 2017;55:27–32. doi: 10.1016/j.neurobiolaging.2017.03.009

78. Skoldunger A, Fastbom J, Wimo A, et al. Impact of Inappropriate Drug Use on Hospitalizations, Mortality, and Costs in Older Persons and Persons with Dementia: Findings from the SNAC Study. Drugs & aging 2015;32(8):671–8. doi: 10.1007/s40266-015-0287-4 [published Online First: 2015/08/02]

79. Pasina L, Lucca U, Tettamanti M. Relation between anticholinergic burden and cognitive impairment: Results from the Monzino 80-plus population-based study. 2020;29(12):1696–702. doi: 10.1002/pds.5159

80. Pfistermeister B, Tümena T, Gaßmann K-G, et al. Anticholinergic burden and cognitive function in a large German cohort of hospitalized geriatric patients. PloS one 2017;12(2):e0171353. doi: 10.1371/journal.pone.0171353

81. Risacher SL, McDonald BC, Tallman EF, et al. Association Between Anticholinergic Medication Use and Cognition, Brain Metabolism, and Brain Atrophy in Cognitively Normal Older Adults. JAMA Neurology 2016;73(6):721–32. doi: 10.1001/jamaneurol.2016.0580

82. Shah RC, Janos AL, Kline JE, et al. Cognitive Decline in Older Persons Initiating Anticholinergic Medications. PloS one 2013;8(5):e64111. doi: 10.1371/journal.pone.0064111

83. Swami S, Cohen RA, Kairalla JA, et al. Anticholinergic Drug Use and Risk to Cognitive Performance in Older Adults with Questionable Cognitive Impairment: A Cross-Sectional Analysis. Drugs & aging 2016;33(11):809–18. doi: 10.1007/s40266-016-0400-3

84. Tsoutsoulas C, Mulsant BH, Kumar S, et al. Anticholinergic Burden and Cognition in Older Patients With Schizophrenia. The Journal of clinical psychiatry 2017;78(9):e1284–e90. doi: 10.4088/JCP.17m11523 [published Online First: 2017/12/01]

85. Weigand AJ, Bondi MW, Thomas KR, et al. Association of anticholinergic medications and AD biomarkers with incidence of MCI among cognitively normal older adults. Neurology 2020;95(16):e2295–e304. doi: 10.1212/wnl.0000000000010643 [published Online First: 2020/09/04]

86. Wu YH, Wang CJ, Hung CH, et al. Association between using medications with anticholinergic properties and short-term cognitive decline among older men: A retrospective cohort study in Taiwan. Geriatr Gerontol Int 2017;17 Suppl 1:57–64. doi: 10.1111/ggi.13032 [published Online First: 2017/04/25]

87. Yarnall AJ, Lawson RA, Duncan GW, et al. Anticholinergic Load: Is there a Cognitive Cost in Early Parkinson’s Disease? Journal of Parkinson’s Disease 2015;5:743–47. doi: 10.3233/JPD-150664

88. Esin E, Ergen A, Cankurtaran M, et al. Influence of antimuscarinic therapy on cognitive functions and quality of life in geriatric patients treated for overactive bladder. Aging & Mental Health 2015;19(3):217–23. doi: 10.1080/13607863.2014.922528

89. Geller EJ, Crane AK, Wells EC, et al. Effect of anticholinergic use for the treatment of overactive bladder on cognitive function in postmenopausal women. Clin Drug Investig 2012;32(10):697–705. doi: 10.2165/11635010-000000000-00000

90. Hampel C, Betz D, Burger M, et al. Solifenacin in the Elderly: Results of an Observational Study Measuring Efficacy, Tolerability and Cognitive Effects. Urol Int 2017;98(3):350–57. doi: 10.1159/000455257 [published Online First: 20170202]

91. Iyer S, Lozo S, Botros C, et al. Cognitive changes in women starting anticholinergic medications for overactive bladder: a prospective study. International Urogynecology Journal 2020;31(12):2653–60. doi: 10.1007/s00192-019-04140-3

92. Wagg A, Khullar V, Marschall-Kehrel D, et al. Flexible-Dose Fesoterodine in Elderly Adults with Overactive Bladder: Results of the Randomized, Double-Blind, Placebo-Controlled Study of Fesoterodine in an Aging Population Trial. J Am Geriatr Soc 2013;61(2):185–93. doi: 10.1111/jgs.12088

93. Welk B, McArthur E. Increased risk of dementia among patients with overactive bladder treated with an anticholinergic medication compared to a beta-3 agonist: a population-based cohort study. BJU Int 2020;126(1):183–90. doi: 10.1111/bju.15040 [published Online First: 2020/03/14]

